# Evaluating Generative Video AI for Standardized Psychiatric Patient Simulation With Graded Hygiene Deterioration

**DOI:** 10.64898/2026.06.22.26356153

**Authors:** Benson Mwangi, Hammza Jabbar Abdl Sattar Hamoudi, Mon-Ju Wu, Andrés Martin, Jair C. Soares, Cesar A. Soutullo

## Abstract

**Introduction:** A clinician’s initial assessment during the mental status examination (MSE) places substantial weight on a patient’s general appearance, grooming, and hygiene. However, the logistical difficulty of producing simulated or standardized patient (SP) videos that systematically manipulate these characteristics limits the development of clinical AI tools and training curricula. This pilot study investigates the technical feasibility of using a video-generation diffusion model to re-animate modified reference images onto driving videos, enabling the creation of diverse patient presentations without the need for repeated filming.

**Methods:** Utilizing an established publicly available dataset, we extracted reference images of three SPs and applied a text-to-image AI model to generate five appearance conditions: the unmodified baseline and four escalating hygiene-deterioration levels: mild, moderate, marked, and severe. We then used the *Wan2.2-Animate-14B animate* video generation AI model to re-animate these modified portraits onto the original driving footage. This factorial design varied several model parameters including; pose retargeting, classifier-free guidance scales, and generation modes, resulting in 180 unique videos. Quality was measured through Fréchet Video Distance (FVD) for distributional fidelity and a physics-aware assessment performed by a multimodal large language model to evaluate physical plausibility.

**Results:** Our analysis yielded two primary observations. First, compositing through replacement-mode achieved significantly higher temporal fidelity than animation-mode (mean FVD 8.6 vs. 19.4; Cohen’s d = 1.84). Second, while distributional fidelity showed a monotonic decline as hygiene perturbation increased (Spearman ρ = 0.48, p < 0.001), physics-aware scores did not follow a similar trend. This pattern is consistent with fine-motor artifacts arising from model-level generative constraints rather than from the severity of the appearance modification alone.

**Conclusions:** These findings demonstrate that generating appearance-modulated clinical video libraries is technically achievable. Nevertheless, the persistence of fine-motor artifacts underscores the necessity of expert human oversight before these materials can be safely deployed in educational and translational settings.

## Introduction

A patient’s general appearance, grooming, and hygiene constitute a primary domain of the mental status examination (MSE) and serve as near immediate clinical indicators for diagnostic formulation and illness monitoring (Norris et al., 2016; Quek et al., 2021; Strauss et al., 2021). This observational assessment is especially important when identifying conditions characterized by profound self-neglect, including the negative symptoms of schizophrenia, severe depressive episodes, and advanced neurocognitive disorders (Quek et al., 2021; Strauss et al., 2021). However, appearance assessments in a MSE remain vulnerable to substantial inter-rater variability, with reported kappa values ranging from 0.47 to 0.80 (Chmielewski et al., 2015; Freedman et al., 2013). This unreliability is driven in large part by the absence of controlled stimulus materials that present the same patient at systematically varied levels of grooming and hygiene. Existing standardized patient (SP) video libraries are inherently static, and producing iterative versions of the same individual across graduated levels of self-care not only demands repeated filming sessions and complex makeup protocols but faces strict physical limitations. For instance, an actor cannot ethically or practically be made to undergo clinically meaningful medication-associated weight change on demand, nor can conventional simulation easily reproduce visible somatic, dermatologic, or motor changes associated with substance use disorders or neurodegenerative illnesses. These clinically informative appearance changes, and their combinations, are simply outside the reach of conventional simulation (Lenouvel et al., 2022; Martin et al., 2020; Xie et al., 2015). The field consequently lacks the benchmark stimuli required for clinician training, diagnostic calibrations, and validation of emerging AI-based clinical assessment tools (Cruz-Gonzalez et al., 2025; Passos et al., 2019).

The rapid maturation of computer vision and digital phenotyping tools makes this shortage increasingly consequential. For example, automated systems can now quantify facial expression dynamics (Ambrosen et al., 2025; Bufano et al., 2023; Cohen et al., 2020), psychomotor disturbances (Abbas et al., 2021; Ambrosen et al., 2025; Boutaleb et al., 2026), and multimodal behavioral signatures that distinguish between psychiatric states such as unipolar and bipolar depression (Zhong et al., 2025). Yet the development of these tools has relied upon uncontrolled clinical footage with idiosyncratic appearance variance. However, algorithm validation demands standardized stimuli with known, incremental changes in appearance, a requirement that existing SP libraries cannot satisfy. In parallel, generative AI is beginning to transform psychiatric education and simulation (Ajluni, 2025; Kilincel et al., 2026; Mwangi et al., 2026; Zhang et al., 2026), and recent work has highlighted its broader potential for digital mental health applications (Schildkrout, 2024; Sezgin and McKay, 2024; Weightman et al., 2025). The central technical objective, which forms the premise of this study is therefore to determine whether generative video models can faithfully re-animate a patient’s video whose appearance has been deliberately and systematically modified for clinical utility.

Latent video diffusion models offer a potential mechanism to address this gap. Given a static reference portrait and a temporal driving video, these models synthesize outputs in which the reference subject performs the exact kinematics of the driving input, effectively functioning as high-fidelity simulators of the visual world (Brooks et al., 2024). A rapidly growing family of architectures now targets the specific task of character animation from a single image. For instance, the magicanimate video animation model introduced a diffusion-based framework with a dedicated appearance encoder and temporal attention module for identity preservation (Xu et al., 2023). Animate Anyone and its successor refined this approach through a ReferenceNet neural network architecture and pose guider that improved control over body and facial movements (Hu et al., 2023). DreamActor-M1 extended these methods to learn motion directly from raw video rather than relying on explicit skeleton extraction, at the cost of reduced fine-grained control (Luo et al., 2025). The VACE model proposed a unified framework handling reference-to-video generation, structural conditioning, and masked editing within a single diffusion transformer backbone (Jiang et al., 2025). More recently, the Wan2.2 family (Wan et al., 2025) introduced Wan2.2-Animate-14B, a 14 billion parameter model purpose built for character animation that unifies both full-frame generation and environment-preserving character replacement within a single jointly trained backbone (Cheng et al., 2025). These reference-to-video animation models share a common paradigm in which a reference image supplies appearance and a driving signal supplies motion, but they differ in how they composite the generated subject into the output frame as well as how faithfully they preserve identity across extended sequences.

Wan2.2-Animate-14B warrants particular attention for potential clinical and training translation. First, it implements two distinct compositing pipelines within the same diffusion backbone. In animation mode, the generated subject is rendered against an artificial neutral background, isolating the character from environmental context. In replacement mode, the model uses instance segmentation masking to extract the subject from the original frame and composites the re-animated subject back into the preserved spatial environment. Second, in recent standardized self-reconstruction benchmarks, Wan-Animate achieved an FVD of 118.65, Structural Similarity Index Measure (SSIM) of 0.813, and Learned Perceptual Image Patch Similarity (LPIPS) of 0.227 on a self-reconstruction character-animation benchmark, surpassing all compared open weight models including StableAnimator (FVD 147.92) and UniAnimate (FVD 155.03). For portrait-level facial animation, it achieved an FVD of 94.65, outperforming X-Portrait2 (FVD 98.03) and SkyReel-A1 (FVD 101.45) (Cheng et al., 2025). This dual-mode design, combined with its state-of-the-art open weight performance, made Wan2.2-Animate-14B the natural choice for the present study.

Translating these models into clinical use, however, introduces real risks. Despite their impressive global realism, generative architectures remain susceptible to visual hallucinations, particularly fine-motor anatomical distortions such as physically implausible kinematics (Aithal et al., 2024; Narasimhaswamy et al., 2024; Soroudi et al., 2025). In psychiatric training and digital phenotyping, such artifacts carry significant clinical implications. For example, a hallucinated tremor could be misinterpreted as psychomotor agitation or a medication-induced movement disorder, corrupting both human learning and algorithmic validation. We therefore benchmarked the fidelity of Wan2.2-Animate-14B in re-animating psychiatric SPs from the (Martin et al., 2020) dataset across four graded levels of clinically relevant hygiene deterioration. Each appearance condition was tested under a full factorial design varying compositing architecture, classifier-free guidance scale, and pose retargeting, yielding 180 generated videos. Fidelity was quantified using Fréchet Video Distance (FVD), and physical plausibility was evaluated across five anatomical and kinematic dimensions by a multimodal large language model (LLM) serving as a physics-aware rater. Together, these analyses establish the first empirical evidence on whether latent video diffusion can produce controlled, appearance-modulated clinical video libraries suitable for psychiatric education, inter-rater calibration, AI benchmarking and generation of synthetic AI model fine tuning datasets.

## Methods

### Study Design

We employed a full factorial design crossing three generative variables of the Wan2.2-Animate-14B model (Cheng et al., 2025). The first variable was compositing architecture or generation model, tested at two levels (animation mode and replacement mode). The second was classifier-free guidance (CFG) scale, tested at three levels (1, 4, and 6). The third was pose retargeting, tested at two levels (enabled and disabled). This 2 × 3 × 2 design produced 12 distinct generation configurations. Each configuration was applied to three SP identities across five appearance conditions (an unmodified original plus mild, moderate, marked, and severe hygiene deterioration), yielding a final analytic sample of 180 generated videos.

Ground-truth clinical materials were drawn from the validated SP video repository developed by (Martin et al., 2020). This dataset features professional actors portraying composite psychiatric phenotypes derived from longitudinal clinical practice, presented across sequential video segments of escalating symptom burden. The repository demonstrates robust inter-rater reliability (κ = 0.66, p < 0.001) when scored against the ABC-STAMPS rubric (Appearance, Behavior, Cooperation, Speech, Thought process and content, Affect, Mood, Perceptions, Suicidality) (Martin et al., 2020). We used the complete cohort of three SP identities available in the repository, each mapped to a distinct clinical phenotype namely; “Ben” - Obsessive compulsive disorder (OCD), “Karthik” - Schizophrenia and “Robbin” - Bipolar Disorder (BD). The driving videos depicted these subjects (Ben, Karthik and Robbin) seated during a simulated clinical interview, during which they exhibited natural conversational speech and accompanying hand gestures (see Supplementary materials).

To generate controlled appearance variation, static reference portraits were extracted from the driving videos and edited using the Google Gemini text-to-image model (Gemini Team et al., 2023; Imagen-Team-Google: Jason Baldridge et al., 2024). A structured prompting protocol (detailed in Supplementary Methods) produced four escalating tiers of hygiene deterioration: mild, moderate, marked, and severe. For analysis, severity was coded as 0 = original, 1 = mild, 2 = moderate, 3 = marked, and 4 = severe. The perturbation targeted the appearance variables central to the MSE, specifically hair condition, facial-hair grooming, and attire degradation. Notably, to isolate appearance as the sole manipulated variable, explicit negative constraints in the prompt prohibited changes to patient identity, somatic geometry, or background. However, it is known that latent video diffusion models are highly sensitive to the spatial resolution of their reference inputs, and low-fidelity images reliably trigger temporal collapse and visual artifacting during motion synthesis (Cheng et al., 2025; Rombach et al., 2021). Therefore, to prevent this phenomena, the gemini generated images were upscaled using the enhanced super-resolution generative adversarial network (ESRGAN) (Rakotonirina and Rasoanaivo, 2020), which restored the high-frequency spatial detail necessary to stabilize the downstream diffusion process.

The super-resolved portraits and their corresponding driving videos were then passed to Wan2.2-Animate-14B, which synthesizes an output video in which the reference subject performs the kinematics of the driving source (See Fig 1). Three generation parameters were varied in the factorial design. Notably, each parameter governs a fundamentally different aspect of the synthesis process and was expected, on architectural grounds, to interact with the degree of appearance perturbation.

**Figure 1.**
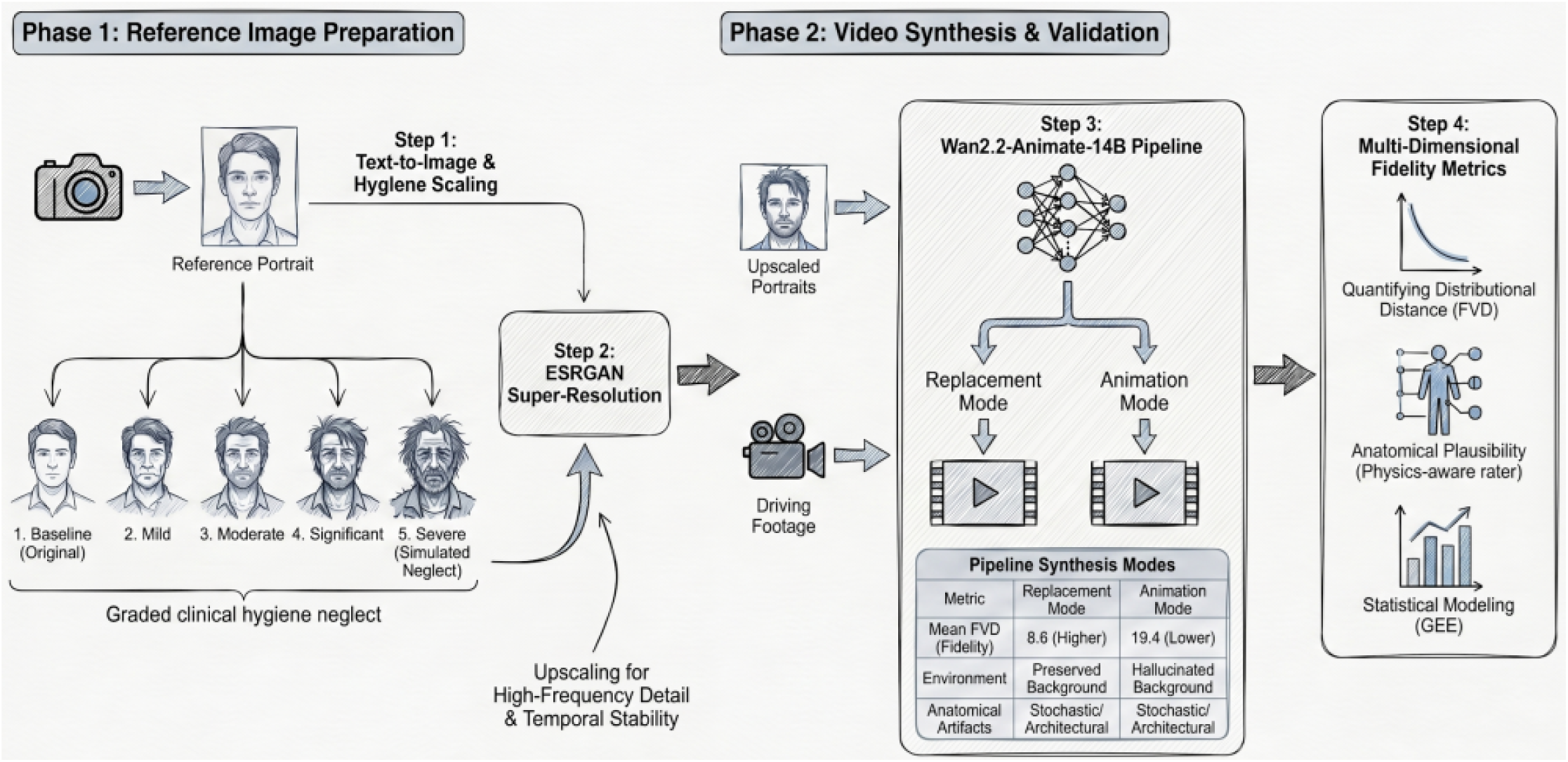
Study pipeline for generating and evaluating appearance.modulated standardized patient videos. In Phase 1, static portraits were extracted from validated SP driving videos (Martin et al. 2020) and modified using Google Gemini text-to-image generation to produce five graded levels of clinical hygiene neglect (original through severe). The resulting images were upscaled through ESRGAN super-resolution to restore the high-frequency spatial detail required for stable downstream diffusion synthesis. In Phase 2, the upscaled portraits and their corresponding driving footage were passed to the Wan2.2-Animate-14B pipeline, which synthesized output videos in two generation modes. Replacement mode regenerates only the masked subject region and composites it into the preserved real background. Animation mode generates both subject and background entirely from the diffusion process.. Each generated video was evaluated along two complementary quality axes, distributional fidelity through Fréchet Video Distance (FVD) and anatomical plausibility through a physics-aware multimodal rater (Qwen3-Omni). Results were analyzed using generalized estimating equations within the full factorial design. The embedded table summarizes performance differences between generation modes. *N* = 180 videos (3 patients × 2 modes × 3 CFG scales × 2 retarget settings × 5 hygiene levels).

The first parameter to be tested was generation mode. The model implements two compositing pipelines within a single jointly trained backbone (Cheng et al., 2025). In animation mode, the target frames are initialized as zero-filled latents, and the model generates both the subject and the background from the diffusion process, producing an image-to-video synthesis in which the background is derived from the reference portrait. However, in replacement mode, the subject in each driving frame is first removed via instance segmentation masking, and the remaining background is passed to the model as a preserved condition. The model therefore generates content only within the masked subject region, compositing the re-animated reference identity back into the original spatial environment. This is followed by a relighting Low-rank adaptation (LoRA) module which further adjusts the character’s lighting and color tone to match the ambient illumination of the target scene. Therefore, because replacement mode retains the real background and constrains the generative solution space to the subject region alone, it was expected to produce outputs whose global visual statistics remain closer to those of the ground-truth driving video footage.

The second tested parameter was the classifier-free guidance (CFG) scale, which controls the trade-off between the model prompt adherence and output diversity during inference. A value of 1 applies no prompt amplification and yields outputs that reflect the model’s learned distribution with maximum diversity. Higher values progressively amplify the influence of the conditioning signal, producing outputs that conform more tightly to the specified content but with increased risk of oversaturation and structural artifacts. We tested values of 1, 4, and 6, spanning unconstrained generation through moderate-to-strong prompt enforcement. The Wan2.2-Animate-14B prompt used in this study is shown in the supplementary materials.

The third examined parameter was pose retargeting. The model extracts a 2D skeleton from the driving video using ViTPose whole-body pose estimation (Xu et al., 2022) and injects this skeleton as a spatially aligned control signal for body motion. Therefore, when the body proportions of the reference subject differ from those of the driving subject, the raw skeleton may introduce some geometric distortions. Pose retargeting addresses this by rescaling the skeleton keypoints to match the reference anatomy before injection. Retargeting is recommended for animation mode when body proportions are dissimilar and is disabled by default in replacement mode to preserve spatial interactions between the character and the environment (Cheng et al., 2025). We tested both states across both generation modes to empirically quantify the effect rather than rely on default settings.

All remaining hyperparameters (random seed, temporal reference frames, diffusion sampling steps, frame rate, numerical precision, and output resolution) were held constant across the 180 generation runs to ensure that observed differences were attributable solely to the three critical hyperparameters examined. The random seed was fixed at 42 for reproducibility. Temporal reference frames were set to 5 (the recommended default for temporal consistency), diffusion sampling steps to 50, and frame rate to 30 fps, matching the model’s native configuration. Precision was set to bfloat16 and output resolution was read dynamically from each driving video to preserve its native dimensions. All generation runs were executed on a single NVIDIA RTX 6000 Blackwell GPU with 96 GB of video memory and output resolution was read dynamically from each driving video to preserve its native dimensions. Lastly, for each of the 180 generation runs, the first 10 seconds of the driving video were animated, yielding a uniform clip duration across all experimental conditions. The 10 seconds duration was chosen to balance clinical representativeness against the substantial computational cost of diffusion-based inference with a 14-billion-parameter model across 180 generation runs.

Distributional distance between each generated video and its source driving counterpart was quantified using a ResNet3D-18-based Fréchet video feature distance (Xiang et al., 2025), reported here as FVD for consistency with video-generation evaluation conventions. This feature-distance metric served as the primary distributional fidelity outcome because it captures divergence between the original and generated videos. Notably, lower FVD values indicate closer similarity between the original or source video and the AI model generated video. All metric extraction and aggregation were performed with the open-source Artificial Intelligence Generative Video Evaluation (AIGVE) toolkit (Mwangi, 2026; Xiang et al., 2025).

While FVD provides a reliable summary of global distributional quality, it is inherently insensitive to localized anatomical anomalies that may be clinically consequential in diffusion-generated video (Ge et al., 2024). A finger hallucination or an implausible movement trajectory will not substantially shift the global feature distribution, yet either artifact could be mistaken for psychomotor tremor, dyskinesia, or medication-induced extrapyramidal signs in a clinical training context. To capture these localized failures, we implemented an automated physics-aware evaluation powered by the Qwen3-Omni multimodal foundation model (Xu et al., 2025), specifically the Qwen3-Omni-30B-A3B-Thinking variant, which supports reasoning-oriented analysis of video input (Mwangi et al., 2026). Specifically, each video was scored at temperature 0.2 on a 5-point ordinal scale ranging from 1 (severe, pervasive structural violation) to 5 (no detectable violation). Additional implementation details are provided in the Supplementary Materials. Five physical dimensions were evaluated, each selected because it probes a failure mode with a distinct clinical consequence. Finger count consistency (S1) detects supernumerary or missing digits that could be misread as fine-motor pathology. Joint movement physics (S2) evaluates whether articulations follow anatomically plausible trajectories, since violations could mimic dyskinetic or extrapyramidal movement. Arm count stability (S4) flags gross limb duplication or disappearance, an artifact that severely compromises clinical utility. Kinetic momentum conservation (S5) assesses whether body-part velocities exhibit physically continuous transitions, as discontinuities could mimic psychomotor agitation or bradykinesia. Finally, shadow consistency (S6) evaluates whether cast shadows align with the character’s pose and the scene’s lighting geometry, an attribute on which replacement-mode environmental realism depends. The survey questions representing S1-S6 are shown in the Supplementary Materials.

### Statistical Analysis

FVD, the sole continuous outcome, was modeled using Generalized Estimating Equations (GEE) with a Gaussian family, identity link, and exchangeable working correlation structure (Zeger and Liang, 1986). The primary inferential question was whether generation mode, CFG scale, retargeting, and hygiene level, individually or in combination, significantly affect the distributional fidelity of the synthesized videos. Patient identity served as the clustering variable to account for the repeated-measures structure in which each of the three subjects contributed 60 correlated observations. The model included the four main effects and all six two-way interactions. Omnibus Wald tests from the GEE model were used as exploratory tests of association. Because only three SP identities contributed repeated observations, sandwich-based GEE inference is unstable and may be anti-conservative. Therefore, GEE p-values are reported descriptively and are not treated as confirmatory evidence.

The five individual survey dimension scores (S1, S2, S4, S5, S6) were treated as ordinal outcomes and analyzed non-parametrically to determine which generation parameters drive each specific category of physical artifact. Kruskal-Wallis H tests assessed omnibus differences across levels of each experimental factor, and Mann-Whitney U tests assessed pairwise comparisons, with Cliff’s delta reported as the effect size. No composite index was used as a primary outcome. Individual dimension scores were retained as the primary artifact outcomes to preserve the clinical interpretability of each failure mode. A descriptive mean physics score was used as a composite and only for variance decomposition and global configuration ranking. Pairwise comparisons for FVD used Welch’s t-test with Cohen’s d and Bonferroni correction was applied to all families of pairwise comparisons throughout.

A secondary question was whether increasing appearance perturbation itself degrades video quality, independent of how the video is generated. This was tested through a dedicated hygiene dose-response analysis combining three complementary approaches. One-way ANOVA tested for omnibus differences in FVD across the five hygiene levels, and Kruskal-Wallis H tests served the same purpose for the five ordinal survey dimensions. We subsequently used the spearman rank correlation to assess whether any observed degradation follows a monotonic trend. This test examined whether the quality of generated videos worsened progressively as hygiene severity increased, using the ordinal severity code (original = 0 through severe = 4). A covariate-adjusted GEE isolated the hygiene-level effect after controlling for mode, CFG scale, and retargeting, establishing whether appearance perturbation affects fidelity above and beyond the generation parameters. Finally, to identify the generation settings that produce the best overall output across all evaluation criteria simultaneously, optimal configurations were determined through combinatorial Borda count ranking and Pareto dominance analysis across the six retained metrics (FVD and the five survey dimensions). All analyses were conducted in Python 3.11 using Scipy and Statsmodels packages (Seabold and Perktold, 2010; Virtanen et al., 2020).

## Ethics Statement

The standardized patient videos used in this study were derived from a peer-reviewed, open-access educational resource published under a Creative Commons Attribution-NonCommercial license (Martin et al., 2020). The original study was approved by the Yale Human Investigations Committee (No. 2000024005). The videos depict professional actors portraying scripted psychiatric presentations and do not contain real patient data. Because these materials are publicly available educational resources and were reused within the terms of their open-access license, no additional consent or re-consent from the standardized patients was required for the present secondary analysis. No new participants were recruited, no intervention or interaction with human subjects occurred, and no private identifiable health information was collected or analyzed.

## Data Availability

The simulated patient video recordings used in this study are available as appendices to the original publication (Martin et al., 2020). The complete prompts are included in the supplementary materials.

## Author Contributions

B.M. conceived the study, developed the AI software pipeline, conducted the statistical analyses, and contributed to manuscript preparation. H.J.A.S.H. contributed to study conception, statistical analysis, and manuscript writing. C.A.S. contributed to study conception, interpretation of the statistical findings, and critical revision of the manuscript. J.C.S. contributed to study conception, interpretation of the statistical findings, and critical revision of the manuscript. M.-J.W. contributed to study conceptualization, data management, and manuscript drafting. A.M. authored the source clinical publication from which the standardized patient materials were derived and contributed to study conception and manuscript drafting.

## Results

### Data Completeness

All 180 generated videos were processed successfully and returned valid FVD values within the expected range. Physics-aware evaluations were obtained across the five retained dimensions (S1, S2, S4, S5, S6) for the full sample. Complete observations were obtained on three dimensions (S1, S4, S6), while joint movement (S2, *n* = 141) and momentum conservation (S5, *n* = 130) returned lower counts because the relevant phenomenon was absent from a subset of sampled frames, yielding assessor responses of “not applicable.” All analyses used available data for each dimension without imputation.

### Generation Mode Dominates Distributional Fidelity

Of the four experimental factors, generation mode exerted by far the strongest influence on video fidelity. Replacement mode produced substantially lower FVD than animation mode (mean 8.6 vs. 19.4), a difference that was significant in the exploratory GEE model (p < 0.001) and large in magnitude (Cohen’s d = 1.84). This result is consistent with the architectural distinction between the two pipelines. Replacement mode preserves the original spatial environment through instance segmentation masking, so the generated frames remain distributionally close to the ground-truth driving footage. Animation mode must instead synthesize an artificial background from the reference portrait, which shifts the global visual statistics of the output and inflates its divergence from the reference. This effect was the single largest in the entire analysis and held consistently across all three patients and all five hygiene conditions (Figure 2A).

**Figure 2.**
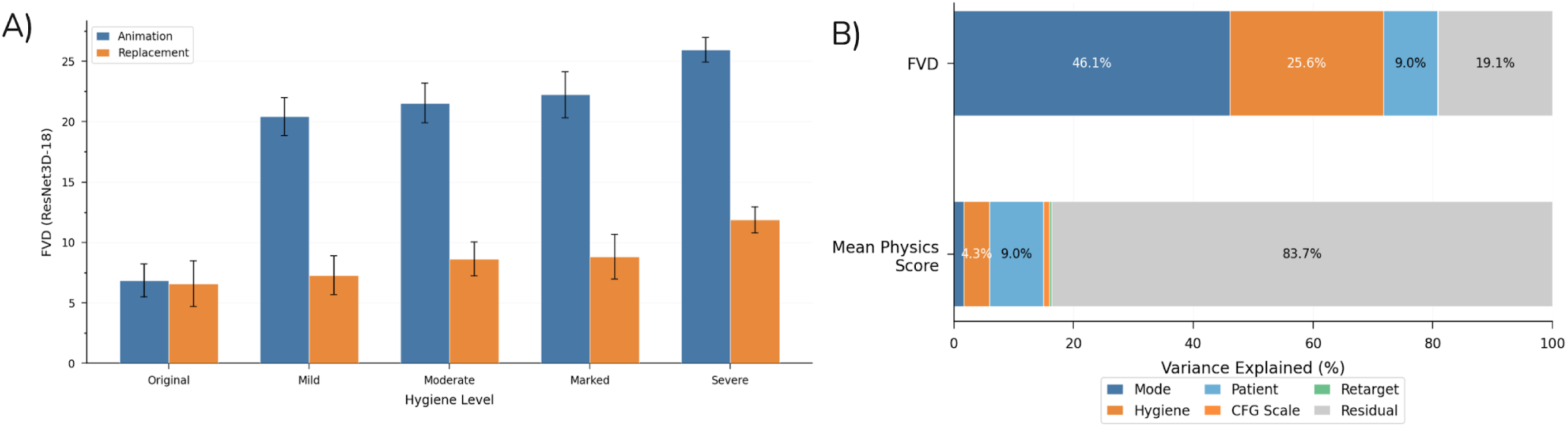
Generation mode dominates distributional fidelity while physics-aware quality remains largely unexplained by experimental factors. **(A)** Mean Fréchet Video Distance (FVD; ResNet3D-18 backbone) by generation mode across five hygiene perturbation levels. Replacement mode produced substantially lower FVD than animation mode at every level (mean 8.6 vs. 19.4; Cohen’s *d* = 1.84, *p* < 0.001). FVD increased monotonically with perturbation severity (Spearman ρ = 0.479, *p* < 0.001). Error bars denote 95% confidence intervals. **(B)** Variance decomposition (η^2^ from factorial ANOVA) for FVD (upper bar) and mean physics score across the five retained dimensions (lower bar). Generation mode (46.1%) and hygiene level (25.6%) together explained over 70% of FVD variance. For the mean physics score, no factor exceeded 9.0% and 83.7% remained residual, indicating that physics artifacts arise from stochastic properties of the diffusion process rather than from the manipulated parameters. *N* = 180 videos (3 patients × 2 modes × 3 CFG scales × 2 retarget settings × 5 hygiene levels).

Variance decomposition confirmed the dominance of generation mode, which accounted for 46.1% of the total FVD variance, followed by hygiene level (25.6%) and patient identity (9.0%). CFG scale and retarget each contributed less than 1%. In contrast, a descriptive variance decomposition applied to the mean physics score showed that no single experimental factor explained more than 15% of the variance, with 76.6% remaining residual (Figure 2B). This asymmetry indicates that distributional fidelity is highly sensitive to the manipulated parameters, whereas physics-aware quality is governed primarily by stochastic properties of the diffusion process.

### Guidance Scale and Pose Retargeting Show Conditional Effects

CFG scale showed a nominal association with FVD in the exploratory GEE model (Wald χ² = 48.54, *df* = 2, *p* < 0.001), but no pairwise contrast between individual CFG levels survived Bonferroni correction, indicating that the effect is detectable in aggregate yet modest in magnitude and entangled with other factors. Pose retargeting had no significant main effect on FVD (Wald χ² = 1.07, *df* = 1, *p* = 0.300). The mode-by-retarget interaction was nominally significant in the exploratory GEE model (Wald χ² = 16.66, *df* = 2, *p* < 0.001), as was the CFG-by-retarget interaction was also nominally significant (Wald χ² = 16.15, *df* = 2, *p* < 0.001), indicating that the influence of retargeting depends on both the compositing architecture and the guidance scale (Figure 4B). These conditional effects argue for evaluating generation parameters jointly rather than in isolation.

### Physical Plausibility Is Largely Invariant to Generation Parameters

In contrast to FVD, the physics-aware survey scores were largely insensitive to the generation parameters. Across mode, CFG scale, retarget, and hygiene level, the only statistically significant omnibus effect among the five dimensions was arm count stability (S4) by generation mode (Kruskal-Wallis *H* = 17.97, *p* < 0.001), with replacement mode scoring higher than animation mode (Cliff’s δ = -0.224, small effect). The pairwise Mann-Whitney *U* test confirmed this difference after Bonferroni correction (*p* < 0.001). No other dimension reached significance for any predictor (Supplementary Figures S1–S4).

This divergence between the evaluation metrics warrants closer examination. FVD captures global spatiotemporal fidelity relative to the ground-truth reference, whereas the physics survey probes localized anatomical coherence at the level of individual fingers, joints, and limbs. That generation parameters move FVD substantially while leaving the physics scores essentially unchanged indicates that parameter tuning operates on the macro-statistical properties of the video rather than on the fine-grained anatomical reasoning that governs finger, joint, and momentum artifacts.

### Hygiene Severity Degrades Distributional Fidelity but Not Physical Plausibility

A dedicated dose-response analysis tested whether increasing appearance perturbation degraded video quality independent of the generation parameters. For distributional fidelity, the relationship was unambiguous (Figure 3A). The omnibus ANOVA was significant (*p* < 0.001), and a Spearman trend test confirmed a significant positive monotonic association between perturbation severity and FVD (ρ = 0.479, *p* < 0.001) (See Supplementary Figure 6). The more severely the reference image was altered, the further the generated video drifted from the ground-truth distribution. The covariate-adjusted GEE confirmed this effect after controlling for mode, CFG scale, and retarget (*p* < 0.001). All four perturbed levels differed significantly from the unperturbed baseline after Bonferroni correction, and the mean divergence grew monotonically (mild Δ = 7.12, Cohen’s *d* = 1.22; moderate Δ = 8.35, *d* = 1.45; marked Δ = 8.80, *d* = 1.44; severe Δ = 12.19, *d* = 2.08; See Table 1).

**Figure 3.**
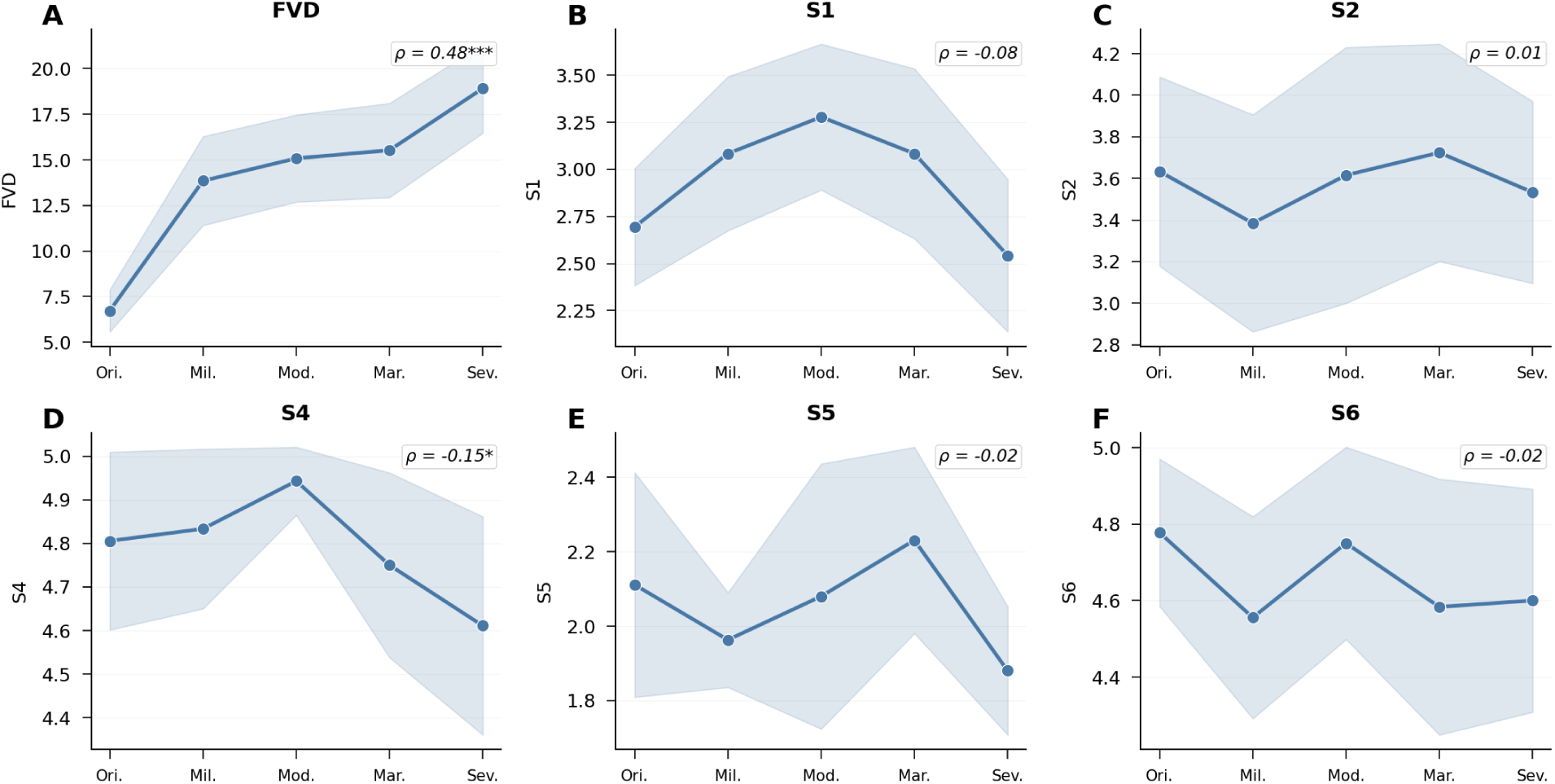
Appearance perturbation severity produces a clear dose-response degradation of distributional fidelity but not of physical plausibility. Mean metric values across five levels of appearance perturbation (original, mild, moderate, marked, severe; *n* = 36 videos per level). Shaded bands indicate 95% confidence intervals. Spearman ρ and significance are annotated in each panel *(*p* < 0.05, *\*\*\*p* < 0.001). **(A)** FVD increased monotonically with perturbation severity (ρ = 0.48, *p* < 0.001), and all four perturbed levels differed significantly from the original baseline after Bonferroni correction (Cohen’s *d* ranging from 1.22 for mild to 2.08 for severe; see Table 1). **(B-F)** The five retained physics dimensions showed no comparable monotonic dose-response. S1 (ρ = -0.08), S2 (ρ = 0.01), S5 (ρ = -0.02), and S6 (ρ = -0.02) showed no significant trend (all *p* > 0.30). S4 produced the only nominally significant correlation (ρ = -0.15, *p* = 0.042), but no pairwise comparison against the original baseline survived Bonferroni correction and the trajectory was non-monotonic, peaking at moderate before declining. This dissociation indicates that physics artifacts such as finger hallucinations, momentum violations, and shadow inconsistencies are intrinsic to the diffusion architecture and not driven by the degree of appearance departure from the original reference.

**Table 1.**
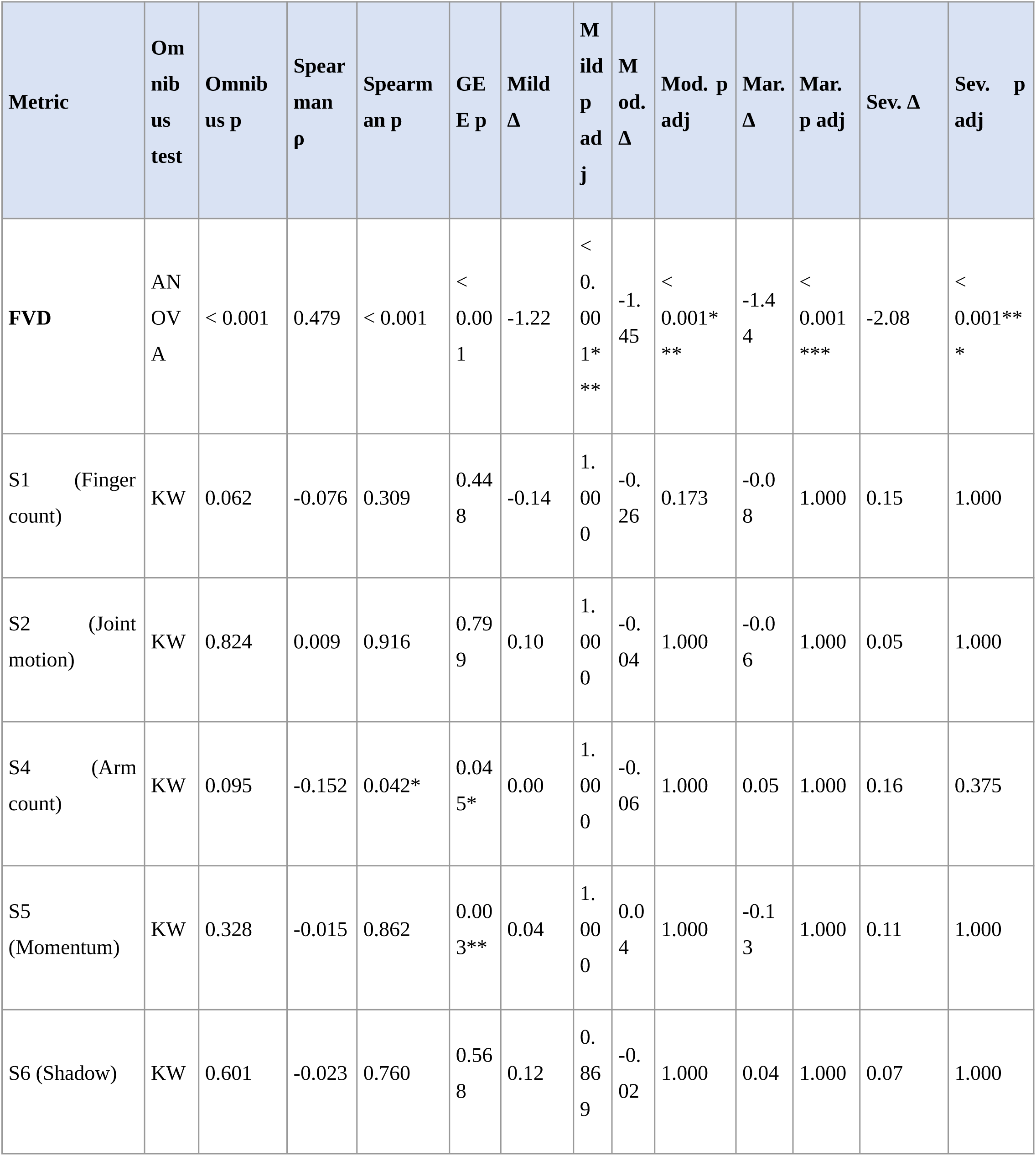
Hygiene dose-response summary. Omnibus tests, Spearman monotonic trend, covariate-adjusted GEE, and pairwise comparisons against the original (unperturbed) baseline for FVD and each retained physics dimension. Pairwise effect sizes are Cohen’s *d* for FVD and Cliff’s δ for survey dimensions. Adjusted *p*-values reflect Bonferroni correction for four comparisons. KW = Kruskal-Wallis. \**p* < 0.05, \*\**p* < 0.01, \*\*\**p* < 0.001.

The physics-aware scores told a different story (Figure 3B-F). Only one of the five retained dimensions produced a nominally significant monotonic trend: arm count (S4, ρ = -0.152, *p* = 0.042), but no pairwise contrast against baseline survived Bonferroni correction and the trajectory was non-monotonic. The remaining dimensions were non-significant across severity levels: finger count (S1, ρ = -0.076, *p* = 0.309), joint movement (S2, ρ = 0.009, *p* = 0.916), momentum (S5, ρ = -0.015, *p* = 0.862), and shadow consistency (S6, ρ = -0.023, *p* = 0.760; See Table 1).

However, although unadjusted monotonic trends were non-significant for the survey dimensions, the covariate-adjusted GEE detected a significant hygiene effect for momentum (S5, *p* = 0.003). Once generation parameters are held constant, hygiene level exerts a small detectable influence on this dimension, but the effect is neither monotonic nor large enough to register in the unadjusted trend test. This pattern is most consistent with subtle, non-systematic perturbation effects rather than the clean dose-response degradation observed for FVD. This dissociation represents a primary finding of the hygiene analysis. The physical artifacts detected by the survey, including finger hallucinations, momentum violations, and shadow inconsistencies, are not driven by how severely the reference image departs from the original. They appear instead to be intrinsic limitations of the diffusion architecture that surface regardless of perturbation severity.

### Cross-Domain Correlation Between Distributional Fidelity and Physical Plausibility

To test whether distributional similarity predicts physical plausibility, Spearman correlations were computed between FVD and each of the five retained survey dimensions. The strongest association was with arm count (S4, ρ = -0.33, *p* < 0.001), indicating that videos with lower FVD tended to preserve correct arm count more reliably. The remaining dimensions showed weak, non-significant correlations with FVD (S2 ρ = -0.14; S5 ρ = -0.10; S1 ρ = -0.04; S6 ρ = 0.01; Supplementary Figures S2, S6). These correlations were weak to moderate, reinforcing the conclusion that distributional fidelity and localized physical coherence capture complementary dimensions of video quality. A video that is distributionally close to its reference is not guaranteed to be anatomically correct, and vice versa.

### Optimal Configuration

Combinatorial Borda count ranking across the six metrics (FVD and the five survey dimensions) identified replacement mode with CFG = 1 and retargeting disabled as the globally optimal configuration (mean FVD = 8.29). Pareto dominance analysis identified seven non-dominated configurations. Five of six replacement-mode configurations were Pareto-optimal, whereas only two animation-mode configurations achieved Pareto status (animation with CFG = 1 and animation with CFG = 6, both with retargeting disabled). No animation-mode configuration with retargeting enabled was Pareto-optimal (See Supplementary Figure 7). Figure 4A shows the best-achievable FVD for each patient across the five hygiene levels; all selected videos used replacement mode, and FVD increased with perturbation severity for every patient.

**Figure 4.**
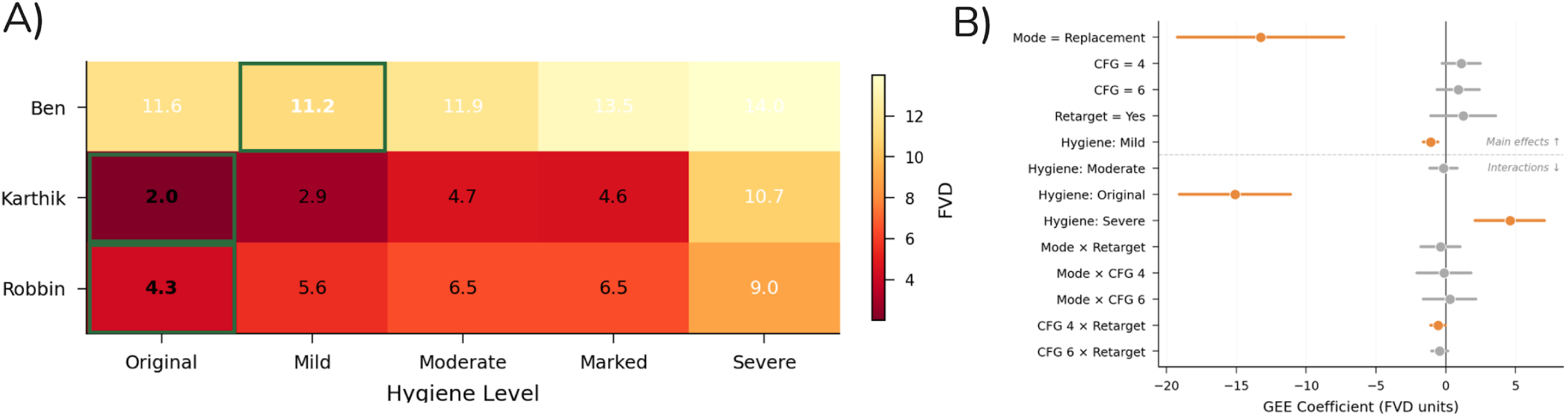
Best-achievable distributional fidelity across patients and perturbation levels, with GEE model coefficients. **(A)** Fréchet Video Distance (FVD) of the best-performing video for each patient at each hygiene level, selected by weighted Borda rank across six evaluation metrics (FVD, S1, S2, S4, S5, S6). All selected videos used replacement mode. Green borders highlight the lowest-FVD cell for each patient. FVD increased monotonically with perturbation severity for all three patients, although the absolute magnitude of degradation varied substantially across patients (Karthik: 2.0 to 10.7; Ben: 11.2 to 14.0; Robbin: 4.3 to 9.0), reflecting differences in the source video characteristics. **(B)** Forest plot of GEE regression coefficients for FVD (exchangeable working correlation; reference categories: animation mode, CFG = 1, retarget = no, marked hygiene). Horizontal bars indicate 95% confidence intervals; orange points denote significant effects (p < 0.05). Replacement mode produced the largest effect (β ≈ -14 FVD units), followed by the original and mild hygiene contrasts. CFG scale and retarget main effects were small and non-significant, though their interactions with mode reached significance (see text). Hygiene interaction terms are omitted due to numerical instability arising from three-cluster GEE estimation (see Methods).

## Discussion

This study provides the first systematic evidence that a latent video diffusion model can re-animate standardized psychiatric patients whose appearance has been deliberately altered across graded levels of hygiene deterioration. Three principal findings emerged. First, replacement-mode compositing produced markedly higher distributional fidelity than animation mode. This effect was large (Cohen’s *d* = 1.84) and consistent across all patients and conditions. Second, distributional fidelity degraded in a clear monotonic dose-response pattern as hygiene perturbation increased (Spearman ρ = 0.479, *p* < 0.001). This establishes a direct link between the degree of reference-image modification and the temporal quality of the generated output. Third, physics-aware quality did not follow the same trajectory. Fine-motor artifacts, including finger hallucinations, implausible joint trajectories, and momentum violations, appeared at comparable rates whether the reference image was unmodified or severely perturbed. This dissociation indicates that localized anatomical artifacts are intrinsic properties of the generative architecture, not byproducts of the appearance-manipulation task.

### Relationship to Prior Work

These findings extend a growing literature on generative AI in psychiatric simulation and medical education (Ajluni, 2025; Martin et al., 2020). Indeed, (Martin et al., 2020) recently demonstrated that curated SP video libraries improve student MSE performance but the static nature of those recordings has remained a persistent constraint. Our approach addresses this constraint by utilizing a single recorded clinical interview as a temporal scaffold onto which an array of appearance variants can be projected. Stimulus production is thereby converted from a laborious recording process into a video generation task and the marginal cost of each new variant drops from days of studio time to minutes of computation. This reframing is particularly important because recent scoping reviews have identified content scalability and standardization as the central unmet needs in simulation-based education (Weightman et al., 2025). Importantly, initial evidence suggests that AI-powered virtual simulated patients can improve clinical interviewing skills in psychopathology training (Eraslan and Thomann, 2025), reinforcing the case for scalable, generative approaches to stimulus production.

Our quantitative findings also corroborate and sharpen qualitative observations from adjacent work. For example, (Chu and Goodell, 2024) reported persistent visual artifacts, including facial distortion and “visual stuttering”, during synthetic-patient video generation for difficult-conversation training. In another study, (Haider et al., 2025) described high patient usability for an AI-generated physician avatar in postoperative education but flagged movement naturalness as a recurring limitation. Our physics-aware evaluation framework converts these anecdotal reports into structured, dimensional measurements that localize exactly which physical domains fail. Crucially, it also demonstrates that these failures occur independently of the global distributional realism that FVD captures.

Notably, the dominance of replacement mode is consistent with the architectural logic described by (Cheng et al., 2025). Animation mode must generate both the subject and the background entirely from the latent diffusion process while on the other hand, replacement mode, regenerates only the masked subject region and composites it back into the preserved real background. This constrained solution space reduces the distributional drift that inevitably accumulates when an entire scene must be creatively generated from scratch. Consequently, the practical implication for a future clinical or training deployment is that replacement mode should serve as the default pipeline.

### Distributional Fidelity and the Role of Fréchet Video Distance

Fréchet-style video feature distances have become widely used for evaluating generative video models since the introduction of FVD by (Unterthiner et al., 2018) which has been widely adopted for benchmarking architectures including CogVideoX, Wan2.2, and Sora (Brooks et al., 2024; Wan et al., 2025). In this study, FVD demonstrated interpretable and consistent behavior by sharply differentiating replacement from animation mode while simultaneously tracking the hygiene dose-response monotonically. Together, these properties support it’s utility as a distributional summary statistic in controlled experimental settings, particularly when the generative and reference distributions share a common visual domain.

Recent work, however, has revealed that FVD is not the comprehensive quality indicator it is often assumed to be. For example, recently (Ge et al., 2024) showed that FVD exhibits a strong content bias toward per-frame spatial quality while remaining surprisingly insensitive to temporal distortions. In addition, (Luo et al., 2024) extended this critique by demonstrating that the I3D feature space underlying FVD violates the Gaussian distributional assumptions on which Fréchet distance depends and that reliable estimation requires impractically large sample sizes. Taken together, these findings establish that FVD captures global distributional fidelity but can miss localized, frame-level anomalies.

Our results confirm this blind spot in a clinical-video context. The strongest correlation between FVD and any physics-aware dimension was with arm count stability (S4, ρ = −0.33). The remaining dimensions showed weak, non-significant associations (S1 ρ = −0.04; S2 ρ = −0.14; S5 ρ = −0.10; S6 ρ = 0.01). A video can therefore score well on distributional fidelity while still containing hallucinated fingers or implausible joint trajectories and the reverse also holds. For clinical training scenarios in which an aberrant tremor could be mistaken for psychomotor pathology, this dissociation makes FVD necessary but insufficient as a standalone quality gate.

### The FVD-Physics Dissociation and Its Architectural Implications

The dissociation between distributional fidelity and physical plausibility has consequences for both evaluation methodology and model development. Variance decomposition revealed that generation mode alone accounted for 46.1% of FVD variance while hygiene level followed at 25.6%, and patient identity contributed 9.0%. By contrast, no single experimental factor explained more than 15% of the variance in physics survey scores and 76.6% of variance remained residual (Figure 2B). This asymmetry indicates that distributional fidelity is highly responsive to the manipulated parameters. Physics-aware quality, however, is governed primarily by stochastic properties of the generative model diffusion sampling process. This interpretation aligns with findings from the broader video-generation evaluation literature. Indeed, interpretability analyses of large-scale video encoders have shown that physics-related and appearance-related representations occupy nearly orthogonal subspaces (Joseph et al., 2026), providing a structural basis for the functional independence observed here. The VideoPhy benchmark (Bansal et al., 2025) established that current text-to-video models achieve physical commonsense scores below 20% across most tested architectures. This reveals a fundamental gap in the ability of generative models to produce physically plausible motion. In another benchmark study, PhyGenBench (Meng et al., 2024) similarly documented widespread violations of physical laws spanning mechanics, optics, and material properties across leading video generation systems. Therefore, our data extend these observations to the clinical-video domain. We found that fine-motor artifacts persist across diverse model architectures and remain perturbation-invariant within individual models, appearing at the same rate whether the system produces a faithful likeness or a severely altered one. This indicates that a model that improves on distributional benchmarks is therefore not guaranteed to produce more anatomically faithful output. Therefore, for high-stakes clinical applications, distributional and physics-aware evaluations must be conducted separately, as overall performance cannot serve as a proxy for anatomical correctness.

### Observations on the Perturbation Pipeline

Two additional observations about the perturbation pipeline merit discussion, although both were qualitative rather than quantitative. First, in our preliminary model screen for generating hygiene-perturbed reference images, the Google Gemini text-to-image model (Gemini Team et al., 2023) produced the most consistent responses to structured prompting for graded appearance modification. Alternative models either failed to preserve patient identity under perturbation, produced appearance changes that did not correspond to the prompted severity level, or ignored the hygiene-related prompt components altogether. Second, even within Google Gemini, the generated severity levels were not perceptually equidistant. The lower levels (mild, moderate) were visually similar to one another and to the original, whereas the upper levels (marked, severe) diverged disproportionately (See supplementary materials). This nonlinear mapping is consistent with known behavior in text-to-image diffusion models, where extreme prompt modifiers exert disproportionate influence on the generated output relative to moderate ones (Sadat et al., 2024). The hygiene perturbation variable in this study should therefore be interpreted as ordinal rather than interval-scaled, and all dose-response analyses treat it accordingly. Noticeably, achieving perceptually calibrated severity gradients will likely require text-to-image model fine-tuning or learned prompt interpolation rather than manual prompt engineering alone.

### Clinical Applications Beyond Hygiene

The monotonic dose-response relationship between hygiene severity and distributional fidelity establishes that the pipeline can produce ordered, graded appearance changes from a single reference image. Our finding that physics-aware artifacts remained perturbation-invariant further implies that the anatomical limitations documented here are properties of the generative model itself, not of the hygiene axis specifically. This invariance is consistent with evidence that physical variables in video models are encoded in high-dimensional, distributed subspaces that are largely decoupled from appearance-level representations (Joseph et al., 2026). Together, these two results suggest that the same replacement-mode pipeline could be applied to other clinically important appearance dimensions that share a common constraint where they cannot be ethically or practically simulated with a live actor. For instance, second-generation antipsychotics, particularly olanzapine and clozapine, produce clinically significant weight gain that can reach 3.0 kg over acute treatment (Pillinger et al., 2020). Recognizing this change matters because medication-induced weight gain is a leading driver of antipsychotic non-adherence and subsequent relapse. Yet a live SP cannot ethically gain or lose substantial weight on demand. The same factorial approach applied here to hygiene could, in principle, benchmark generative weight-change trajectories against distributional and physics-aware thresholds before deployment. As another example, the physical signs of anorexia nervosa, including emaciation, temporal wasting, lanugo, and cachexia, are diagnostically central yet impossible to simulate safely with a live actor. Patients also frequently conceal emaciation under loose or layered clothing (Treasure et al., 2011), a subtle presentation that is difficult to teach without controlled examples. Generative modification could produce graded depictions of progressive restriction as well as the weight-restoration phase of treatment, spanning a clinical spectrum that is currently absent from standardized teaching materials. Chronic alcohol, methamphetamine, and opioid use each produce characteristic patterns of facial and somatic change (Basu et al., 2018). Experienced clinicians learn to recognize these informally, but they are rarely taught as explicit assessment targets. Controlled renderings of substance-related decline, from the dermatologic and dental sequelae of stimulant use to the stigmata of chronic liver disease, would convert incidental pattern recognition into a structured curriculum objective. Lastly, the appearance changes of advanced dementia, including diminished grooming, postural change, and reduced eye contact, present a similar simulation challenge. Generative video could depict the appearance trajectory of cognitive decline in a single synthetic patient over modeled time. This would give trainees a longitudinal perspective that cross-sectional clinical exposure rarely affords.

Each of the proposed clinical extensions would inherit the same evaluation architecture validated here. Replacement-mode generation would serve as the default pipeline given its distributional superiority (Cohen’s *d* = 1.84). FVD would quantify global fidelity against the source footage. And the physics-aware survey would screen for the fine-motor artifacts that FVD cannot detect. Across these extensions, only the perturbation axis changes; the factorial benchmarking workflow and the dual-metric evaluation framework that our results establish as necessary for clinical-grade quality assurance remain constant.

### Broader Implications for Digital Phenotyping and AI Benchmarking

The findings reported above extend beyond psychiatric education into four additional application domains where the combination of factorial appearance control and dual-metric evaluation addresses existing methodological gaps.

### Adversarial testing for digital phenotyping

The dissociation between FVD and physical realism presents a direct challenge for AI classifiers trained to detect psychomotor signs. Fine-motor artifacts in our data were perturbation-invariant and detached from global distributional quality. Validating phenotyping systems solely against distributional metrics therefore leaves them blind to anatomical distortions that can mimic clinical pathology. The factorial framework described here offers a corrective where, studies can generate appearance-modulated synthetic libraries in which the perturbation axis and severity level are both known, and use these as adversarial test beds to verify that classifiers remain robust when a patient’s appearance is progressively altered by medication, substance use, or disease progression.

### Standardized stimuli for inter-rater calibration

The monotonic dose-response relationship between hygiene severity and FVD (Spearman ρ = 0.479) suggests a second application. By rendering a single synthetic patient across known, ordered severity levels, educators can create calibration stimulus sets analogous to quality-assurance phantoms in radiology. Training programs could then measure how well trainees and residents converge in their ratings at each severity tier and monitor calibration drift longitudinally. The optimal configuration identified here (replacement mode, CFG = 1, retargeting disabled) provides a ready blueprint for generating these standardized assets at scale.

### Ethically scalable data sharing

Sharing identifiable patient footage for research and training is heavily restricted by privacy and regulatory constraints. Prior work has demonstrated the use of video motion transfer to de-identify faces while preserving diagnostically relevant motor patterns (Cai et al., 2025). Our approach extends this logic in a different direction. Because the generated videos depict synthetically modified SP actors rather than real patients, they reduce some re-identification risks associated with genuine clinical footage. Therefore, under appropriate institutional controls, this pipeline could support broader sharing of educational and algorithmic-development materials while lowering the privacy barriers that currently limit collaborative research.

### Synthetic training data for multimodal video models

The rapid advancement of video-language models and multimodal clinical AI systems is constrained by the scarcity of accessible, well-annotated clinical video data. Our framework addresses this bottleneck directly. Because the factorial design provides precise control over both patient appearance and clinical severity, it can serve as a scalable engine for generating synthetic training datasets with built-in ground-truth labels. Rather than relying on limited real-world cohorts, studies can simulate diverse clinical encounters, including rare or underrepresented presentations, and use these to train or benchmark multimodal models before deployment on genuine patient data.

## Limitations

Several limitations bound these conclusions. First, the study used only three SP identities. This small sample constrains generalizability and limits the GEE to three clusters. The extreme Wald statistics observed in several hygiene interaction terms are direct artifacts of this cluster count and the resulting instability of the sandwich variance estimator. All GEE interaction results are therefore treated as preliminary, and primary hygiene inference is instead routed through the non-parametric dose-response analyses, which do not depend on cluster-count assumptions. Second, artifact evaluation was adjudicated entirely by a multimodal large language model (Qwen3-Omni) rather than by trained human raters. Our implementation used chain-of-thought reasoning and low-temperature sampling to maximize scoring consistency, and the approach provides standardized, scalable preliminary screening. It should not, however, be interpreted as validated clinical or perceptual ground truth. The emerging literature on vision-language models as automated judges has identified persistent challenges, including position bias, verbosity bias, and limited alignment with human perceptual judgments, particularly for fine-grained physical reasoning tasks (Bansal et al., 2025). Convergent validation, in which senior psychiatrists and the automated survey independently rate the same videos, is a necessary next step before these physics scores can be treated as definitive. Third, FVD itself carries known limitations as a distributional metric. Recent work has demonstrated that FVD is biased toward per-frame spatial quality over temporal consistency (Ge et al., 2024), and its underlying I3D feature space violates the Gaussian assumptions on which Fréchet distance depends (Luo et al., 2024). FVD nonetheless produced interpretable, reproducible, and directionally consistent results across all experimental conditions. Its behavior aligned with architectural expectations, consistently yielding lower values for replacement mode than for animation mode. These observations support its practical utility as a relative comparison metric, even though absolute FVD values should be interpreted with caution. Fourth, the perturbation axis was limited to hygiene. The broader applications outlined above, including weight change, substance-related decline, and neurodegenerative progression, are plausible extensions, but their generative fidelity remains to be tested empirically. Finally, the videos were evaluated in isolation rather than embedded in a clinical training exercise. Whether these stimuli actually improve trainee MSE performance therefore awaits a downstream educational trial.

## Future Directions

The most immediate priority is a human-validation study in which senior psychiatry residents and attending physicians rate both the clinical plausibility and the educational utility of the generated videos. This study would directly test whether the artifacts flagged by the automated survey are perceptible and clinically consequential, while simultaneously providing the convergent validation needed to establish multimodal model-based physics scoring as a reliable proxy for expert judgment. Expanding the SP cohort well beyond three identities is equally important, both to enable stable GEE estimation and to examine potential moderators such as skin tone, body habitus, and baseline motion complexity. Applying the framework to alternative perturbation axes, beginning with medication-induced weight change as the most directly quantifiable, would establish the breadth of the approach.

Given the rapid pace of improvement in video diffusion architectures, the fine-motor artifacts documented here may diminish with successive and superior model generations. Longitudinal re-benchmarking, in which each major model release is evaluated against the same factorial design and physics dimensions, would provide a direct empirical record of whether distributional gains translate into anatomical improvements. Such a program would also clarify the timeline on which these models become clinically deployable without human post-production review.

The present findings establish that appearance-modulated clinical video generation is technically feasible and quantitatively tractable. They also provide the evaluation framework, the optimal configuration parameters, and the baseline measurements against which future models can be judged. What remains is to close the loop by placing these videos in front of clinicians, measuring whether they learn from them, and determining where the boundary falls between generative artifact and clinical utility.

## Data Availability

All data used in this study are publicly available under this publication; Martin, A., Jacobs, A., Krause, R., & Amsalem, D. (2020). The mental status exam: an online teaching exercise using video-based depictions by simulated patients. MedEdPORTAL, 16, 10947.

https://www.mededportal.org/doi/abs/10.15766/mep_2374-8265.10947

## Acknowledgments

This work was supported by the NECMHR01 grant from the Texas Child Mental Health Care Consortium (TCMHCC) and The National Institute of Health (NIH) AIM-AHEAD consortium.

## Notes

### Competing Interest Statement

The authors have declared no competing interest.

### Author Declarations

We used standardized or simulated patients' data originally from this paper; Martin, A., Jacobs, A., Krause, R., & Amsalem, D. (2020). The mental status exam: an online teaching exercise using video-based depictions by simulated patients. MedEdPORTAL, 16, 10947. Data used are publicly available.

