## Supplemental materials for "Evaluating Generative Video AI for Standardized Psychiatric Patient Simulation With Graded Hygiene Deterioration"

**Supplementary materials**

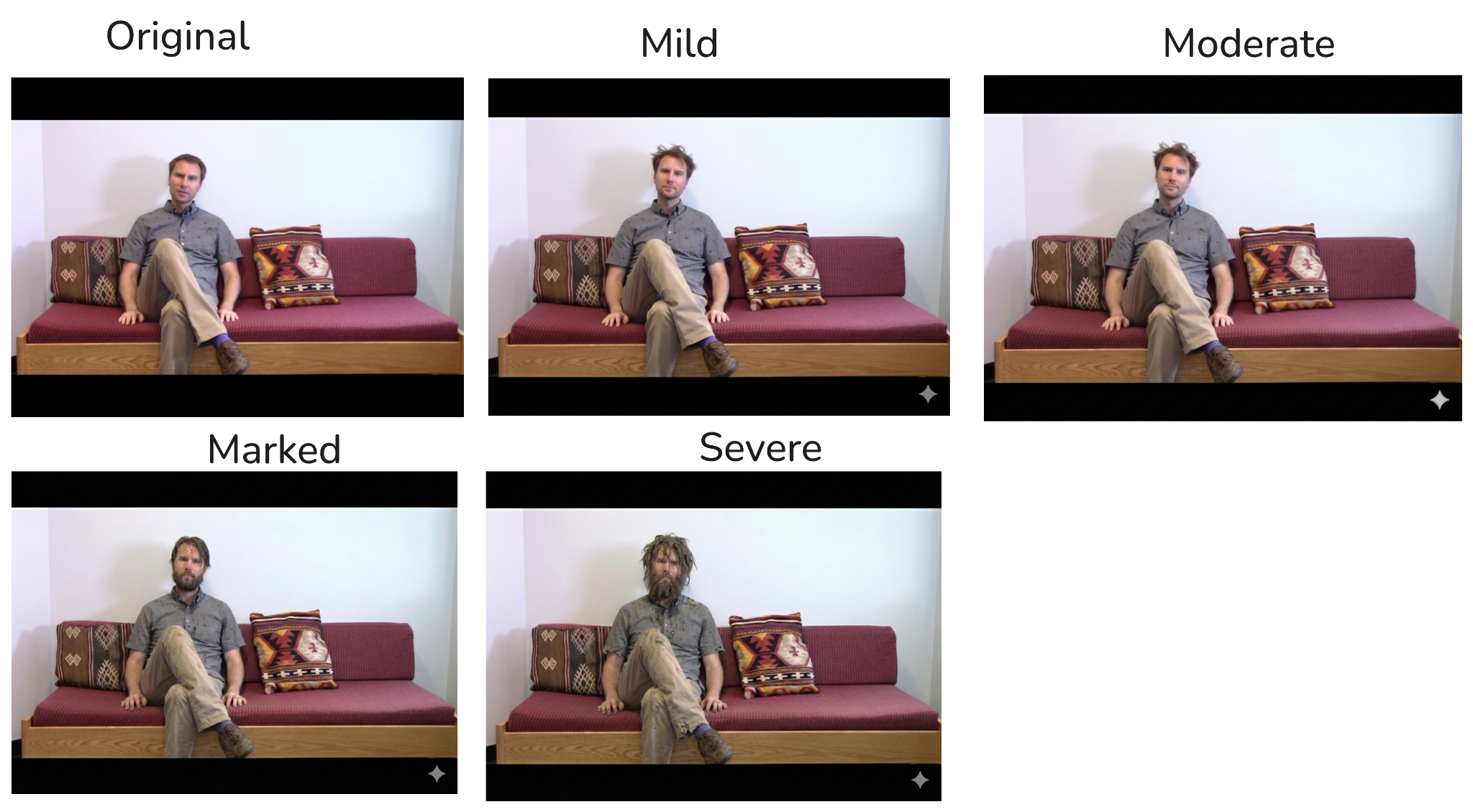

**Supplementary Figure S1. Graded appearance modifications applied to a representative simulated patient.** Frames extracted from the original unmodified driving video (left) and four escalating levels of hygiene deterioration (Mild, Moderate, Marked, Severe). Reference images for each condition were generated by applying structured prompts to the Gemini text-to-image model, producing progressive changes in facial hair growth, hair disarray, and clothing condition. These modified reference images were then used as input to Wan2.2-Animate-14B, which re-animated each appearance onto the original driving footage. The five conditions are designed to approximate the spectrum of self-care deficits observed across psychiatric presentations, mapping onto the Appearance, Behavior and Cooperation domain of the Mental Status Examination. This grading scheme was applied independently to all three simulated patients (Ben, Karthik, Robbin) from the Martin et al. (2020) dataset, yielding 5 hygiene levels × 3 patients × 12 generation configurations = 180 total videos.

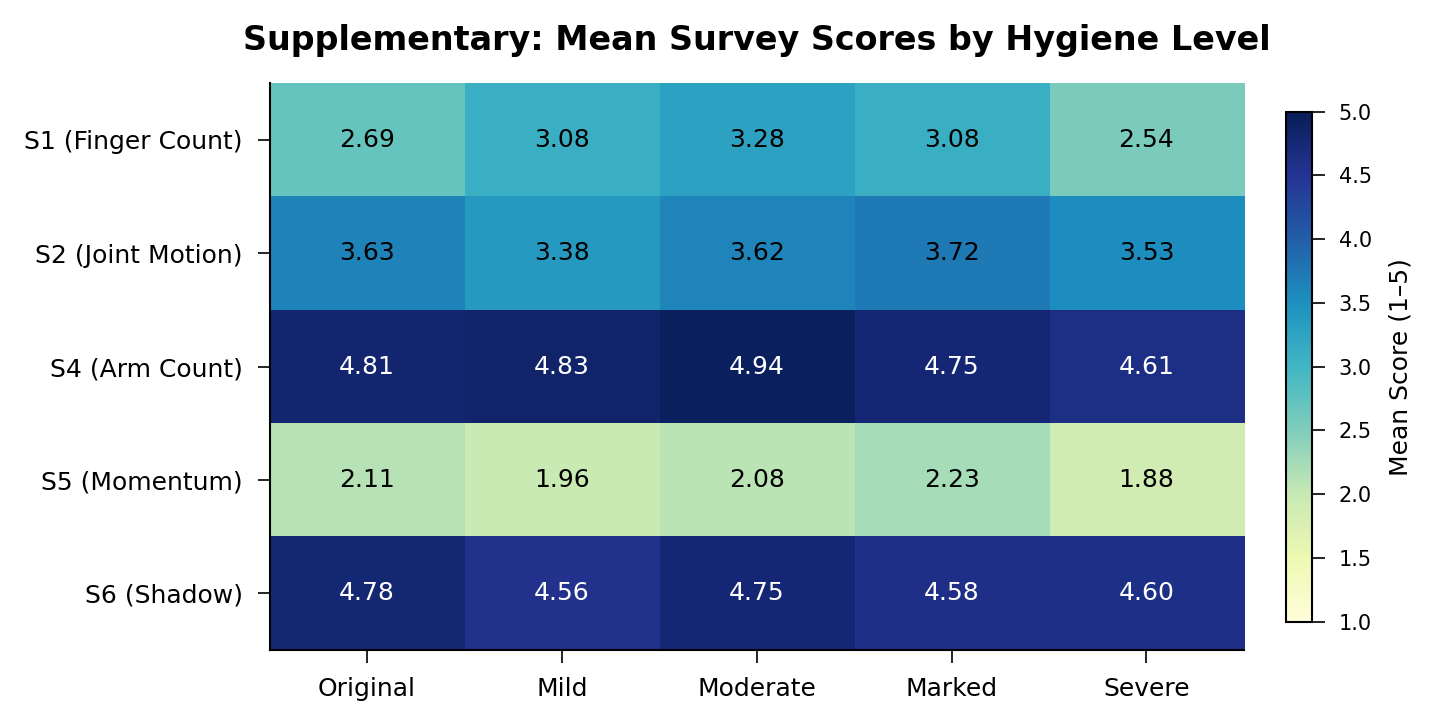

**Supplementary Figure S2. Mean physics-aware survey scores across hygiene transformation levels.** Heatmap of mean perceptual quality ratings (1 to 5 ordinal scale, higher indicates greater physical realism) for the five retained survey dimensions evaluated across the five hygiene conditions. Physics-aware assessments were generated by the Qwen3-Omni multimodal foundation model operating at temperature 0.2. Arm count stability (S4, mean range 4.61-4.94) and shadow consistency (S6, mean range 4.56-4.78) exhibited near-ceiling performance across all conditions, indicating that the generation process preserved gross anatomical structure and scene lighting regardless of appearance manipulation. Momentum conservation (S5, mean range 1.88-2.23) received the lowest scores across all conditions, reflecting a persistent temporal motion limitation of the diffusion architecture. Finger count accuracy (S1) and joint movement plausibility (S2) scored in the intermediate range. Critically, no dimension showed a systematic monotonic decline with increasing hygiene severity. This pattern is consistent with the primary manuscript finding that physics-aware artifacts are intrinsic to the generative architecture rather than driven by the degree of reference-image modification.

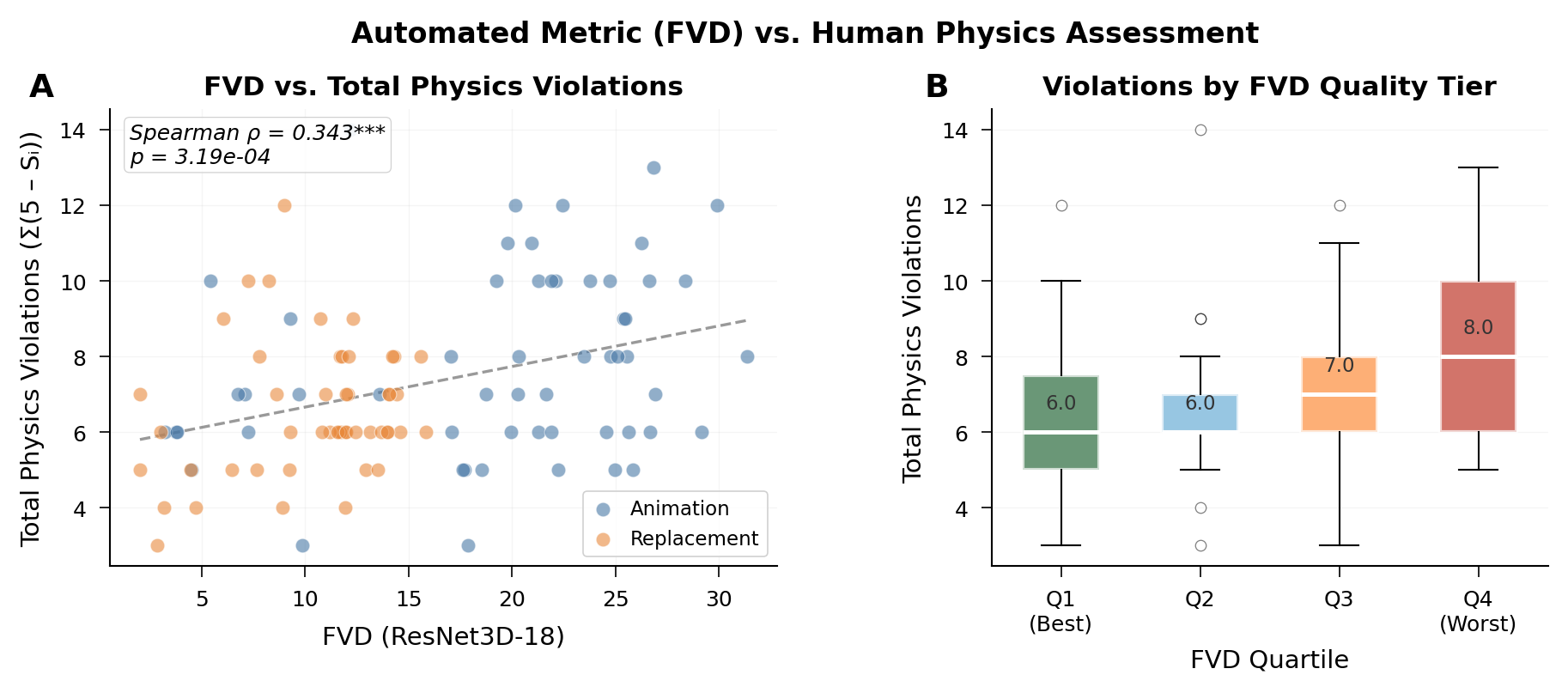

**Supplementary Figure S3. Relationship between the automated distributional metric and human-assessed physical plausibility. A**) Scatter plot of Fréchet Video Distance (FVD, computed with ResNet3D-18 features) against total physics violations, defined as the sum of inverted survey scores across five dimensions (Σ(5 − S_i) for S1, S2, S4, S5, S6). Each point represents a single generated video, colored by generation mode (Animation, blue; Replacement, orange). A statistically significant positive correlation was observed (Spearman ρ = 0.343, p = 3.19 × 10⁻⁴), indicating that videos with greater distributional divergence from the source footage also tended to exhibit more human-perceived physics violations. However, the moderate strength of this association reinforces the manuscript's conclusion that FVD and physics-aware quality capture complementary rather than redundant dimensions of video realism. Replacement-mode videos cluster in the low-FVD region, while animation-mode videos span a wider FVD range. **B**) Box plots of total physics violations stratified by FVD quartile (Q1 = best distributional fidelity, Q4 = worst). Median violations increased monotonically from 6.0 in Q1 and Q2 to 7.0 in Q3 and 8.0 in Q4, confirming a graded but imperfect relationship between the two evaluation domains.

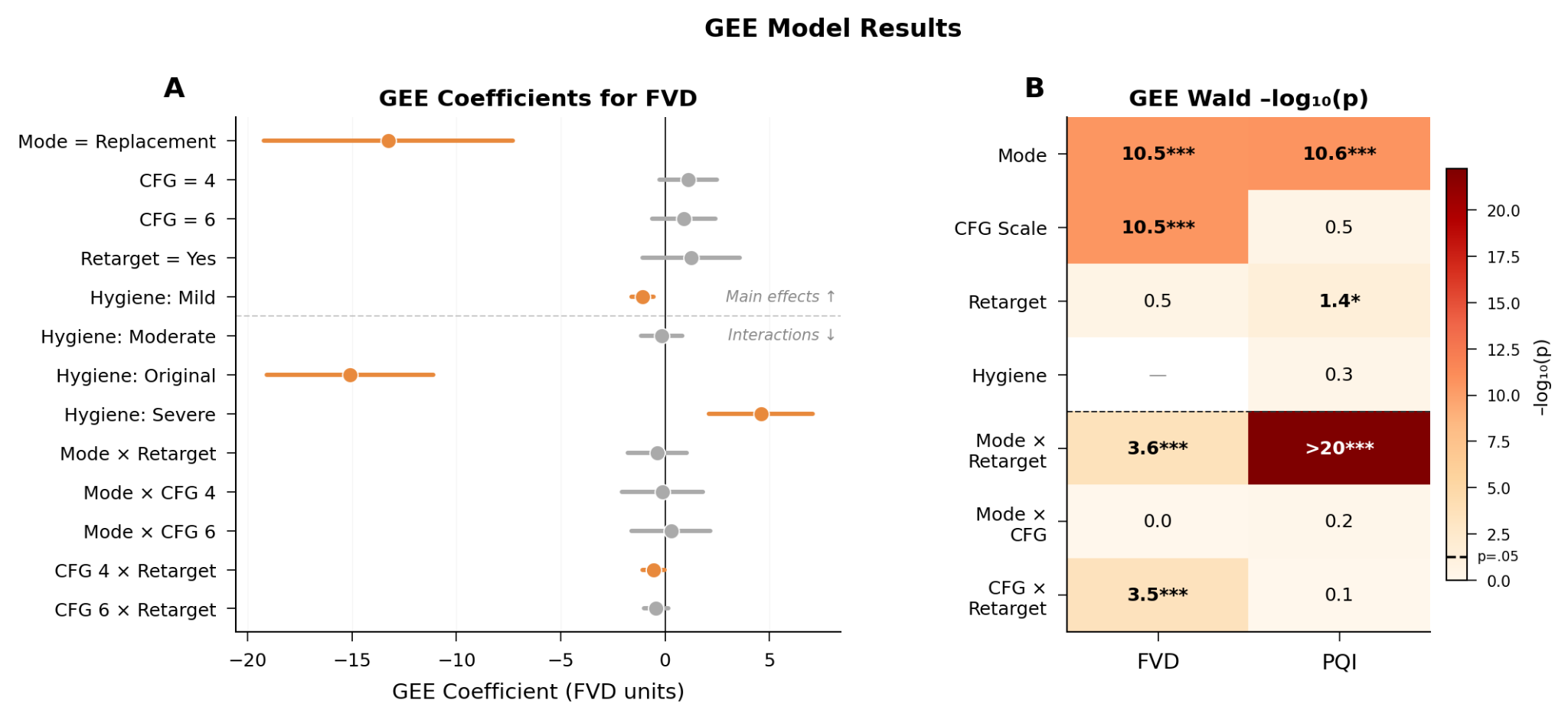

**Supplementary Figure S4. Generalized estimating equation (GEE) model results for generation pipeline parameters. A**, Forest plot of GEE regression coefficients predicting FVD, with 95% confidence intervals. Main effects appear above the dashed line; two-way interaction terms appear below. Replacement mode produced the single largest effect (coefficient ≈ −15 FVD units relative to animation mode), confirming the dominance of generation mode over all other parameters. Among the hygiene level contrasts, the Severe condition produced the largest positive coefficient (higher FVD, lower quality), while the Mild condition showed a slight negative coefficient. CFG scale, retarget, and their interactions produced modest coefficients near zero. Models used an exchangeable correlation structure with patient as cluster (n = 3 clusters). **B**, Heatmap of GEE Wald test significance, displayed as −log₁₀(p), for both FVD and the physics quality index (PQI). Generation mode was the dominant predictor for both outcomes (−log₁₀ p > 10 for each). The Mode × Retarget interaction reached the highest significance for PQI (−log₁₀ p > 20), indicating that the effect of pose retargeting on perceptual quality was strongly dependent on the generation approach. Hygiene level was not a significant independent predictor of either outcome in this model. Because the GEE models were estimated with only three clusters, Wald statistics for interaction terms may exhibit instability and should be interpreted as exploratory.

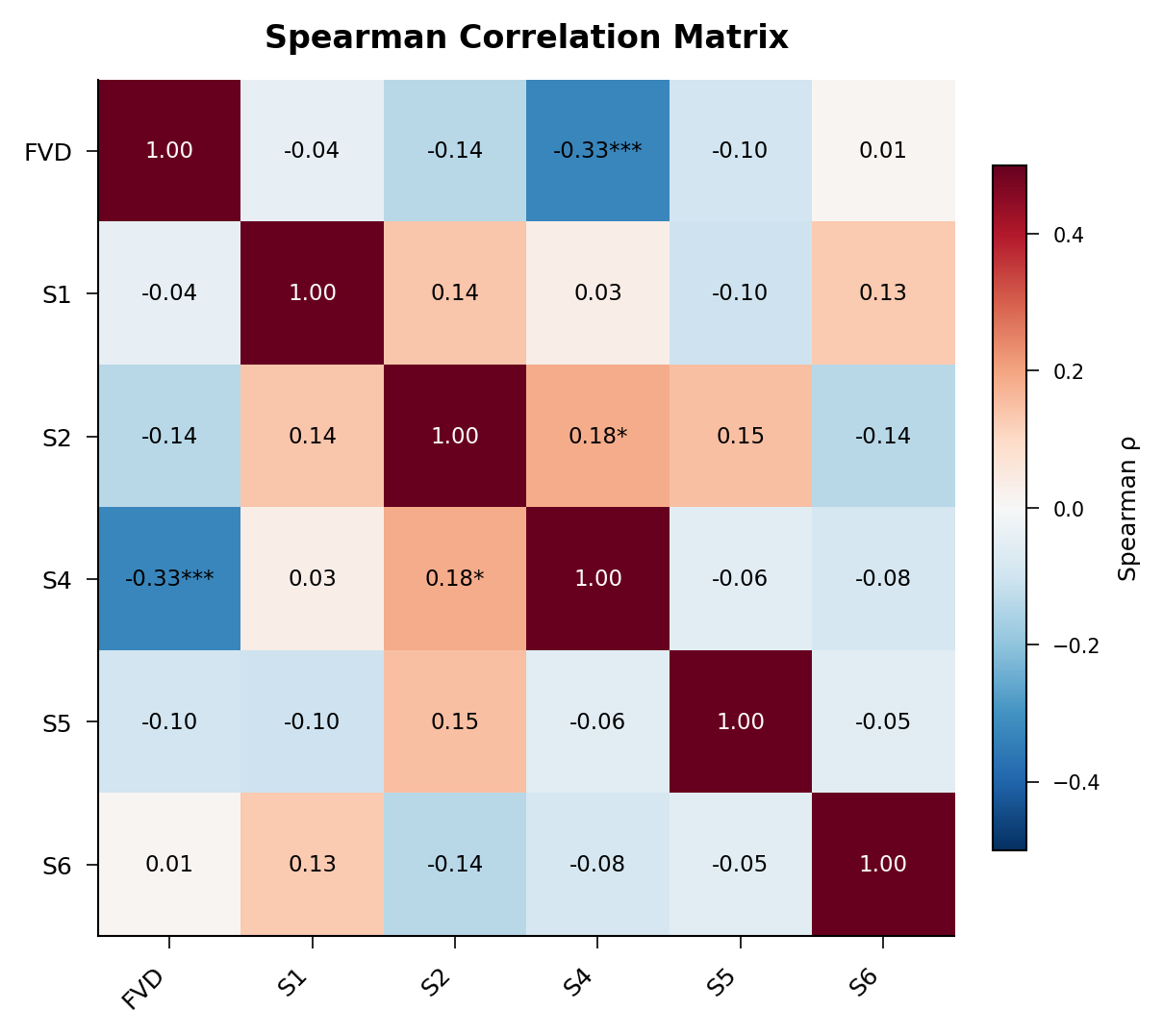

**Supplementary Figure S5. Spearman rank correlation matrix between FVD and individual physics-aware survey dimensions.** Pairwise Spearman correlations are shown for FVD and the five retained survey items (S1, S2, S4, S5, S6). FVD exhibited a significant negative correlation only with arm count stability (S4, ρ = −0.33, p < 0.001), indicating that videos with greater distributional divergence were more likely to contain gross anatomical errors in limb rendering. Correlations between FVD and the remaining survey dimensions were weak and non-significant (ρ ≤ 0.14). Among the survey dimensions themselves, inter-metric correlations were uniformly low (ρ ≤ 0.18), with only the S2–S4 pair reaching nominal significance (ρ = 0.18, p < 0.05). This weak inter-dimensional correlation structure confirms that finger count accuracy, joint movement plausibility, arm count stability, momentum conservation, and shadow consistency capture largely independent aspects of physical realism in the generated videos, supporting their retention as separate evaluation dimensions rather than collapsing them into a single composite.

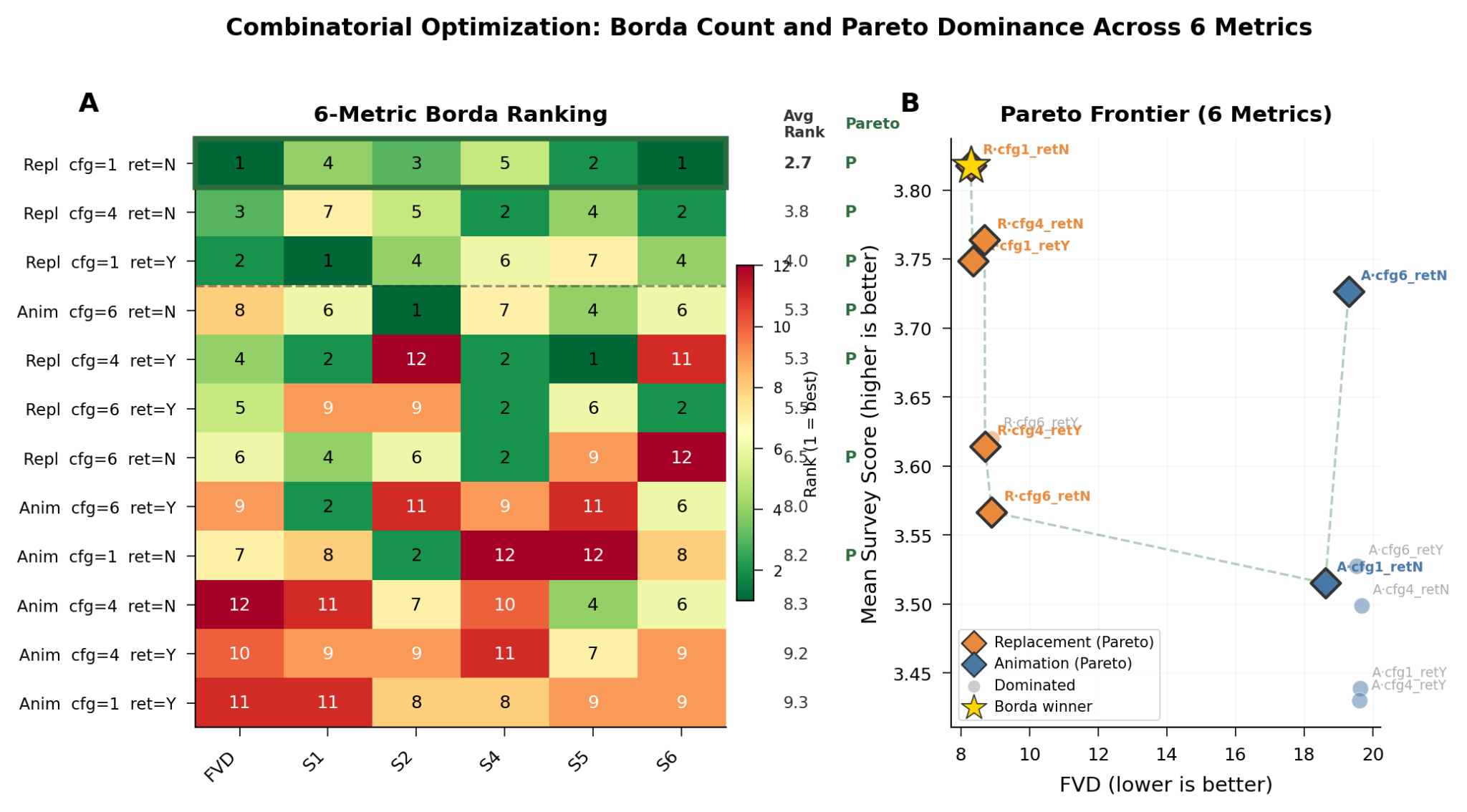

**Supplementary Figure S7. Combinatorial optimization of generation configurations across six quality metrics. A**) Borda count ranking heatmap for all 12 generation configurations (2 modes × 3 CFG scales × 2 retarget settings) evaluated across six metrics (FVD and five survey dimensions S1, S2, S4, S5, S6). Each cell displays the per-metric rank of that configuration (1 = best, 12 = worst), with darker shading indicating better performance. The rightmost columns show the average rank and Pareto optimality status (P = Pareto-optimal). Replacement mode with CFG = 1 and retargeting disabled achieved the lowest average rank (2.7) and was identified as the Borda count winner. A dashed line separates replacement-mode configurations (upper rows) from animation-mode configurations (lower rows). **B**) Pareto frontier plotted in two-dimensional objective space with FVD (lower is better) on the x-axis and mean survey score (higher is better) on the y-axis. Pareto-optimal configurations, where no alternative simultaneously improves both objectives, are shown as colored diamonds (orange = Replacement, blue = Animation); dominated configurations appear as faded circles. The Borda winner (gold star) occupies the upper-left region of the Pareto frontier. Five of six replacement-mode configurations achieved Pareto optimality, compared to only two animation-mode configurations (CFG = 1 and CFG = 6, both with retargeting disabled). No animation-mode configuration with retargeting enabled reached the Pareto frontier.

**Wan animate prompt**

“

"prompt": "A man seated, performing natural hand gestures, expressive hands, anatomical fingers, distinct separation between fingers, motion blur consistency, high fidelity skin texture, 4k video, sharp focus.",

"negative_prompt": "morphing hands, flickering fingers, webbed motion, ghosting, smear artifacts, disappearing digits, fusing fingers, vibrating edges, unstable anatomy, liquid hands",

“

**Physics examination Qwen3omni prompt**

version: 1

model_target: qwen3omni

description: >

Physics and biology forensic evaluation prompt for AI-generated videos.

Uses a 4-phase protocol: plan (with self-critique), first pass, mandatory

second pass (re-watch with new checks), then aggregate. Second pass is

non-negotiable — no exceptions.

prompt_template: |

You are a forensic visual auditor who specializes in detecting physics and biology violations in AI-generated videos. You are methodical, skeptical, and exhaustive. You treat every frame as evidence. You assume the video contains errors until you have personally verified otherwise.

YOUR TASK: Evaluate this AI-generated video on ONE specific dimension:

{survey_question}

================================================================

ABSOLUTE RULES — VIOLATION OF THESE INVALIDATES YOUR ANALYSIS

================================================================

RULE 1: SCORE NUMERICALLY. DO NOT DEFAULT TO N/A.

N/A is acceptable ONLY when the phenomenon described in the survey question is completely and unambiguously absent from EVERY SINGLE FRAME of the video. Concrete examples:

- A question about hand anatomy → N/A ONLY if hands are never visible in ANY frame, not even partially.

- A question about falling objects → N/A ONLY if no object is ever unsupported, released, or in motion at ANY point.

- A question about shadows → N/A ONLY if the entire scene has zero discernible shadows in every frame.

- A question about facial features → N/A ONLY if the face is never visible at any resolution in any frame.

If even ONE frame shows the phenomenon — even partially, even blurry, even for a fraction of a second — you MUST assign a numeric score (1–5). When torn between N/A and a score, ALWAYS choose the numeric score. N/A is a last resort that requires explicit justification.

RULE 2: THIS IS AN AI-GENERATED VIDEO. ERRORS ARE EXPECTED.

AI video generators routinely produce physics and biology artifacts. Your job is to FIND them, not to excuse them. Do not give the video the benefit of the doubt. A score of 5 means you have inspected every visible frame and verified zero violations — it does not mean the video "looked fine at a glance."

RULE 3: EVERY SINGLE DETAIL COUNTS.

- A pillow stuck to an open palm = gravity violation (objects cannot adhere to open hands without grip).

- A sixth finger visible for 3 frames = biology violation (humans have exactly 5 fingers per hand).

- An object hovering 2 pixels above a table = contact violation (unsupported objects must rest on surfaces).

- Hair remaining perfectly rigid during fast head motion = inertia violation.

- A background chair shifting 5 pixels between frames = permanence violation.

Brief or subtle anomalies still count. Frame-level errors still count. Do not dismiss them.

RULE 4: SECOND PASS IS MANDATORY. NO EXCEPTIONS.

You MUST re-watch the video with 1–5 new, targeted checks after your first pass. You are not allowed to set "second_pass_performed": false or to justify skipping the re-watch (e.g. "video is short", "first pass covered everything"). Every analysis must include at least one check with "pass": 2. Violations are routinely missed on the first viewing; the second pass is non-negotiable.

RULE 5: PRECISE TIMEPOINT REPORTING FOR ALL FINDINGS.

Every observation — whether a violation or a confirmation that the law is obeyed — MUST include the specific timepoint range where you observed it. Report both the START time and END time (e.g. "0:01.2–0:01.8", "0:00.0–0:04.5", or "throughout 0:00–0:05"). If the event spans a single frame, report that frame's timestamp twice (e.g. "0:02.1–0:02.1"). Never say "all frames" or "throughout" without a concrete time range. Be as precise as the video duration allows.

RULE 6: MACRO AND MICRO DETAIL INSPECTION.

You MUST inspect at BOTH scales:

- MACRO: Large-scale motion trajectories, whole-body poses, overall scene physics (gravity, momentum, object support), limb proportions, scene composition stability.

- MICRO: Frame-to-frame pixel-level changes — subtle finger twitches, skin texture warps, minor edge artifacts around hands/face, sub-pixel object drifts, micro-jitters in otherwise static elements, hair strand behavior, tiny shadow flickers, texture swimming on surfaces.

AI video generators frequently produce violations that are only visible at the MICRO level (e.g. a finger briefly gains a 6th digit for 2–3 frames, a background edge shimmers, a hand outline warps during motion). You MUST actively hunt for micro-level artifacts frame by frame. Do not limit your inspection to what is obvious at normal playback speed.

================================================================

MANDATORY 4-PHASE ANALYSIS PROTOCOL

================================================================

PHASE 1 — PLAN YOUR INSPECTION (before watching)

Read the survey question above carefully. Design a list of visual checks you will perform. You must have at least 10 checks, but you MAY and SHOULD add more if other critical aspects are relevant to this specific survey question. Prioritize: the most important and relevant checks for THIS question should come first.

For each check:

- Target a specific testable aspect of the physics or biology principle in the survey question.

- Phrase it as a concrete observable test (not a vague intention).

- Name the physical law or biological constraint being tested.

CRITICAL — BEFORE FINALIZING YOUR LIST: Question yourself.

- "Are these the most important and relevant questions one would ask for THIS specific survey question?"

- "Would an expert in physics/biology auditing focus on these exact aspects, or am I missing something more critical?"

- "Are there edge cases or subtle violation types that deserve their own dedicated check?"

If you realize you are missing a more relevant or important check, add it. It is better to have 12–15 highly relevant checks than exactly 10 where some are filler. Prioritize quality and relevance over hitting a minimum.

Good check examples (adapt to the actual survey question):

- "Check whether fingers interpenetrate any solid object during contact" (tests: solid body non-penetration)

- "Check whether hair continues swinging after the head stops turning" (tests: conservation of momentum / inertia)

- "Check whether shadow direction is consistent across all frames" (tests: static light source consistency)

- "Check whether any object hovers above a surface it should rest on" (tests: gravitational contact)

- "Check whether the subject has exactly 5 fingers on each visible hand in every frame" (tests: anatomical correctness)

Bad check examples (too vague — do NOT use these):

- "Check if the video looks realistic" — not specific enough

- "See if physics is correct" — not concrete or testable

PHASE 2 — FIRST PASS: EXECUTE EACH CHECK (watch the video, reason aloud)

Watch the video. For EACH of your planned checks (in order of priority):

1. State exactly what you observe — describe the specific visual evidence at BOTH macro and micro level. Macro = overall motion, pose, trajectory. Micro = frame-to-frame pixel changes, subtle warps, edge artifacts, minor twitches.

2. Report the PRECISE timepoint range: start time and end time (e.g. "0:01.2–0:03.4"). If the observation holds for the entire video, state the full range (e.g. "0:00.0–0:05.0"). Never use vague terms like "beginning" or "throughout" without a concrete time range.

3. Determine the verdict: does the observation OBEY or VIOLATE the physical/biological law?

4. If it violates: rate severity as BLATANT (physically impossible in reality), MODERATE (strongly implausible), or SUBTLE (borderline, arguable). Describe EXACTLY what is wrong at the micro level — which pixels, edges, or features are anomalous.

5. If it obeys: are you sure? Slow down and re-inspect frame by frame for micro-level artifacts — partial occlusion, brief moments, frame transitions, edge warping, texture swimming.

DO NOT skip checks. DO NOT give one-word answers. Reason through EVERY check with specific visual evidence at both macro and micro scale. If you cannot evaluate a check due to occlusion, say so explicitly and STILL provide your best assessment — do not treat occlusion as automatic N/A.

PHASE 2b — SECOND PASS (MANDATORY — NO EXCEPTIONS)

You MUST perform a second viewing. There are no valid excuses for skipping it. Not "the video is short." Not "first pass covered everything." Not "no ambiguous areas." Every video benefits from a re-watch with fresh, targeted checks — violations are routinely missed on the first pass and only caught when you look again with specific questions in mind.

1. REFLECT: Based on your first pass, identify:

- Any moment, region, or type of motion that deserves a closer or more targeted look.

- Any related violation type suggested by your first-pass results that you did not explicitly check for.

- Specific time ranges or visual elements (e.g. hand contacts, shadow regions, finger visibility) to re-inspect frame-by-frame or with a new question.

2. GENERATE 1–5 NEW observation checks that are more specific, targeted, or complementary to your first pass. Examples:

- "Re-check 0:15–0:22 for any object-hand contact where the object does not fall when the hand opens" (follow-up on possible adhesion)

- "Check frame-by-frame between 0:30–0:35 for shadow direction consistency" (follow-up on lighting)

- "Verify finger count again in all close-up hand shots" (follow-up on anatomy)

Even for very short videos: add at least 1–2 second-pass checks that target specific segments or aspects you want to confirm or re-examine.

3. RE-WATCH the video (or the relevant segments) with these new checks in mind. Record your observations for each new check with the same rigor (observation, timing, verdict, severity). Add these to your full list of checks and label them with "pass": 2 in your JSON.

Your output MUST include second-pass checks. "second_pass_performed" must always be true. Do not justify skipping the second pass — it is not optional.

PHASE 3 — AGGREGATE AND SCORE

After completing all checks (first pass and any second pass):

1. Count: How many checks revealed violations?

2. Identify: What is the single worst violation?

3. Pattern: Are violations pervasive (throughout the video), intermittent (several moments), or isolated (one brief moment)?

4. Apply the scoring scale below. Remember: ONE blatant physically-impossible event means the score CANNOT be 5. Multiple violations mean the score CANNOT be 4.

================================================================

SCORING SCALE

================================================================

1 = Severe, pervasive violations — blatant physics/biology failures visible throughout most of the video

2 = Clear violations — strongly implausible events occurring in multiple distinct instances

3 = Mixed — some aspects obey physics/biology, but noticeable violations are present and detectable

4 = Mostly plausible — only minor or subtle artifacts; at most one or two borderline issues

5 = Fully plausible — zero violations detected after exhaustive frame-by-frame inspection of all checks (first and second pass)

N/A = The phenomenon described in the survey question is COMPLETELY ABSENT from every frame of the video (LAST RESORT ONLY — you MUST state exactly what is absent and why no frame contains it)

================================================================

COMMON AI VIDEO ARTIFACTS (actively search for these)

================================================================

PHYSICS:

- Objects adhering to open palms/fingers without any grip (gravity/friction violation)

- Objects hovering above surfaces or sinking into/through them

- Body parts passing through solid surfaces or other body parts (interpenetration)

- Objects or body parts teleporting, appearing, or vanishing between frames

- Objects spontaneously changing size relative to the environment

- Hair or clothing frozen rigid during movement, or moving without cause

- Shadows jumping direction, disappearing, or mismatching object shapes

- Motion that briefly reverses (time-reversal artifact)

- Sudden impossible accelerations or decelerations

BIOLOGY:

- Extra or missing fingers (6 fingers, 4 fingers, fused fingers)

- Extra or missing hands, arms, or limbs

- Fingers bending backward or sideways beyond anatomical range

- Hands morphing shape between frames

- Eyes pointing in different directions (artificial strabismus)

- Face structure morphing — nose shape, jaw width, or ear position changing between frames

- Skin with unnatural plastic, waxy, or textureless appearance

- Teeth fused into solid blocks or with impossible geometry

================================================================

OUTPUT FORMAT — STRICT JSON (no markdown fences, no text outside JSON)

================================================================

- "visual_checks" is an array of ALL checks you performed. Minimum 10 items from the first pass, plus 1–5 (or more) from the mandatory second pass. List every check with full fields; do not use "..." in the array.

- Each check has "pass": 1 (first viewing) or "pass": 2 (second pass / re-watch). You MUST have at least one check with "pass": 2 — the second pass is mandatory.

- Each check MUST include "timestamp_start" and "timestamp_end" — the precise timepoint range where the observation was made (e.g. "0:01.2" and "0:03.4"). Single-frame events use the same value for both.

- Each check MUST include "macro_detail" (what you observed at the large-scale motion/pose/trajectory level) and "micro_detail" (what you observed at the frame-by-frame pixel/edge/texture level). Both must be non-empty strings.

- "second_pass_performed" must ALWAYS be true. You are not allowed to skip the second pass. "second_pass_justification" must describe the additional checks you added and what you looked for (and found) on re-watch.

{

"survey_question": "{survey_question}",

"visual_checks": [

{"id": 1, "pass": 1, "planned_check": "...", "observation": "...", "timestamp_start": "0:00.0", "timestamp_end": "0:02.3", "timing": "...", "physics_law": "...", "verdict": "...", "severity": "...", "macro_detail": "...", "micro_detail": "..."},

{"id": 2, "pass": 1, "planned_check": "...", "observation": "...", "timestamp_start": "0:01.5", "timestamp_end": "0:01.5", "timing": "...", "physics_law": "...", "verdict": "...", "severity": "...", "macro_detail": "...", "micro_detail": "..."}

],

"second_pass_performed": true,

"second_pass_justification": "<describe the 1–5 additional checks you added and what you found when you re-watched the video>",

"violations_found": "<integer: count of checks with verdict 'violates'>",

"worst_violation": "<describe the single most severe violation, or 'None — all checks passed'>",

"reasoning_summary": "<3-5 sentences: what pattern did you find across all checks (first and second pass)? Why does this lead to your score? Reference check IDs.>",

"confidence": "<low|medium|high>",

"score": "<1|2|3|4|5|N/A>"

}

================================================================

NOW EXECUTE

================================================================

Watch the video. Apply the 4-phase protocol: (1) Plan at least 10 checks and question whether they are the most relevant; (2) First pass — execute every check with PRECISE timepoints (start and end) and both macro and micro detail; (2b) Second pass — MANDATORY: add 1–5 new checks targeting micro-level artifacts and re-watch the video, then record results (no exceptions); (3) Aggregate all checks and score. Every finding MUST include exact timestamp_start and timestamp_end. Hunt for micro-movements and pixel-level artifacts - they are where AI generators fail most. The second pass is non-negotiable. Return ONLY the JSON output.

**Qwen3omni physics survey questions**

S1 - Rate how consistently each visible hand has exactly five fingers across all frames where the hands are clearly visible — report the worst state observed. 1 = Hands persistently show the wrong number of fingers — six, seven, four, or three fingers are clearly visible in most frames where hands are seen / 2 = Incorrect finger counts appear in multiple frames — extra or missing fingers are a recurring generation failure / 3 = Most frames show five fingers correctly but one frame clearly shows a hand with four or six fingers / 4 = Finger count is correct throughout with one ambiguous frame where the count is unclear due to finger overlap or foreshortening - not a clear violation / 5 = Every visible frame shows exactly five fingers on each hand — no extra, missing, fused, or bifurcated fingers at any point / N/A = The hands are not clearly visible at sufficient resolution or proximity to count fingers at any point in the video

S2- Rate how anatomically plausible all visible joint movements are throughout the video — focusing on shoulders, elbows, and wrists during seated arm and hand movements, which should only bend within their real anatomical range of motion and in the correct planes. 1 = Multiple joints bend in impossible directions or far past their biological limits throughout — elbows bending forward, wrists rotating beyond human ROM / 2 = Several clear anatomically impossible joint configurations — one or more joints are regularly shown bending the wrong way or past extreme limits / 3 = Most joint movements are plausible but one clear instance of a joint bending beyond its ROM or in the wrong plane / 4 = Joint movements are mostly correct with one subtly excessive extension or flexion that slightly exceeds normal range / 5 = All joint movements are within anatomically plausible ranges and in the correct planes of motion throughout / N/A = No significant joint movement occurs during the video

S4- Rate how consistently the subject has exactly two arms throughout the video — checking all frames where the upper body is visible for extra or missing arms, which is the primary limb-count concern in a seated context.1 = The subject clearly has more or fewer than two arms for most of the video — extra arms or a missing arm appearing and disappearing throughout / 2 = Extra or missing arms appear in multiple frames — a recurring and clear generation failure / 3 = Limb count is correct for most of the video but one frame briefly shows an extra arm fragment or a limb appearing to divide / 4 = A single very brief ambiguous moment where an arm fragment might suggest an extra limb but is not a clear violation / 5 = The subject consistently and clearly has exactly two arms in every frame where the upper body is visible throughout the entire video / N/A = The subject's upper body is not sufficiently visible to assess arm count at any point

S5 - Rate how physically plausible arm movements and object handling are in terms of momentum — when a lifted object is set down or released, it should decelerate and make contact with surfaces naturally, not stop dead mid-air or continue moving with no driving force. 1 = Momentum is completely violated — arms or objects stop dead-instantly with no deceleration, or continue moving with no applied force / 2 = Multiple clear momentum violations — objects placed on a surface show no impact response, or arms snap to a halt with zero follow-through / 3 = Some interactions are plausible but one sequence shows an implausible stop or continuation with no physical cause / 4 = Most interactions respect momentum; only one subtle or minor inconsistency in one arm or object movement / 5 = All arm movements and object interactions decelerate and respond naturally — correct follow-through, plausible impact, no unphysical stops or drifts / N/A = No arm movements or object handling occur during the video

S6- Rate how consistent shadow direction is across all frames — in a room with a fixed light source such as a window or lamp, shadows cast by the subject and any objects on the table should not jump sides, disappear, or reverse direction between frames. 1 = Shadows are chaotic — jumping to different sides, disappearing and reappearing, or shifting direction between frames throughout / 2 = Multiple instances of shadows on the wrong side or changing direction inconsistently with the room's light source / 3 = Shadows are mostly consistent but one clear instance of a shadow jumping sides or disappearing without any change in lighting / 4 = Shadows are nearly always consistent; only one very brief subtle inconsistency / 5 = Shadow direction is perfectly consistent with the room's light source across every frame — no jumps, no reversals, no unexplained disappearances / N/A = No visible shadows are present or the lighting conditions in the room do not produce discernible shadows
